# Personalized modeling of neurodegeneration determines dementia severity from EEG recordings

**DOI:** 10.1101/2023.11.06.23298149

**Authors:** L.G. Amato, A. A. Vergani, M. Lassi, C. Fabbiani, S. Mazzeo, R. Burali, B. Nacmias, S. Sorbi, R. Mannella, A. Grippo, V. Bessi, A. Mazzoni

## Abstract

**INTRODUCTION:** Early identification of dementia is necessary for a timely onset of therapeutic care. However, cortical structural alterations associated with early dementia are difficult to disclose. METHODS: We developed a cortical model of dementia-related neurodegeneration accounting for slowing of local dynamics and global connectivity degradation. We collected EEG recordings at rest from subjects in healthy (HC), Subjective Cognitive Decline (SCD), and Mild Cognitive Impairment (MCI) condition. For each patient, we estimated neurodegeneration model parameters based on individual EEG recordings. RESULTS: Our model outperformed standard EEG analysis not only in discriminating between HC and MCI conditions (F1 score 0.95 vs 0.85) but also in identifying SCD patients with biological hallmarks of Alzheimer’s disease in the cerebrospinal fluid (recall 0.87 vs 0.50). DISCUSSION: Personalized neurodegeneration models could both support classification of MCI and assess the risk of progression from SCD to Alzheimer based only on economical and non-invasive EEG recording

**ClinicalTrials.gov Identifier:** NCT05569083

## Background

Alzheimer’s disease (AD) is the major form of dementia, with more than 10 million people worldwide suffering from it [1]. This number is expected to rise to 50 million in 2050 [2], and several steps ahead in AD research are needed to face this challenge. The disease develops a distinct symptomatology only years or decades after its initial stages, with a progressive decline in memory and other functions. Early diagnosis of AD is therefore a key goal in addressing the progression of the disorder. In the past years a growing attention has been paid to the prodromal and preclinical phases of AD. A prodromal phase is the Subjective Cognitive Decline condition (SCD) [3], in which the subject reports a self-concerned experience of reduced cognitive function, while maintaining normal scores on standardized cognitive tests. In the Mild Cognitive Impairment condition (MCI) [4] affection on one’s cognition is detectable with standardized tests (see [4] and Methods), but there are no direct effects on everyday life. These conditions, although linked to an increase in probability of transition to overt AD for subjects experiencing them [3], are a weak proxy of the disease, with many people suffering from them without ever transitioning to overt AD. In particular, SCD subjects are a very diverse group in both evolution and manifestation of the disease, with little prediction power about progression to AD given by the SCD diagnosis alone [3]. Nowadays, some diagnostic power about dementia progression has been achieved capturing the structural and functional alterations due to the disease with MRI and PET scans [5, 6], or the cerebrospinal fluid (CSF) profiles [7]. However, these procedures are either expensive for the medical facility or discomforting and/or painful for the patient [8]. Evaluation of prodromal and preclinical phases of AD through EEG, which is an affordable and non-invasive procedure, is currently limited to quantifying EEG features known to be biomarkers of the disease [9, 10], such as spectral features and functional connectivity (FC) between brain regions [11, 12]. Despite some interesting results [13], EEG biomarkers fall far behind those obtained from imaging techniques such as MRI and PET scans [14].

The approach that we propose to overcome this issue is that of the personalized brain models [15]. We present a computational framework in which a network simulating cortical activity evolves from healthy condition toward dementia, progressively degenerating local synaptic and global connectivity parameters. In this way we can estimate the EEG features associated with the different stages of this degeneration. Conversely, knowing the specific EEG features of a given patient we can reconstruct the local synaptic and global connectivity parameters and hence evaluate the stage of the patient in the evolution toward dementia. Briefly, we proceeded as follows (Fig. 1). First, we extracted standard biomarkers from experimental EEG recordings (spectral content and functional connectivity, Fig. 1 top row), and we assessed the ability of machine learning algorithms based on these features to discriminate between the patient groups (Fig. 1, top right). Then we developed a dementia progression model starting from The Virtual Brain (TVB) platform [16,17,18,19] framework. We developed a network model in which dementia progression is accounted for by two parameters describing local micro-circuital and large-scale structural alterations of the network caused by AD [20] (Fig. 1 bottom left). The model was validated by comparing the simulated EEG features with population data in the three groups. Subject-specific values of local and large-scale network features were then inferred from the individual values of the aforementioned EEG features (Fig. 1 bottom middle). We assessed then the diagnostic efficacy of these network features in discriminating between patient groups (Fig. 1 Bottom right) and we compared it with the performance of the EEG features. Finally, we compared the efficacy of EEG features and network features in assessing the likelihood of progression from SCD to AD using CSF biomarkers as a proxy [7] and by predicting outcome of the 1y follow-up.

**Fig. 1:**
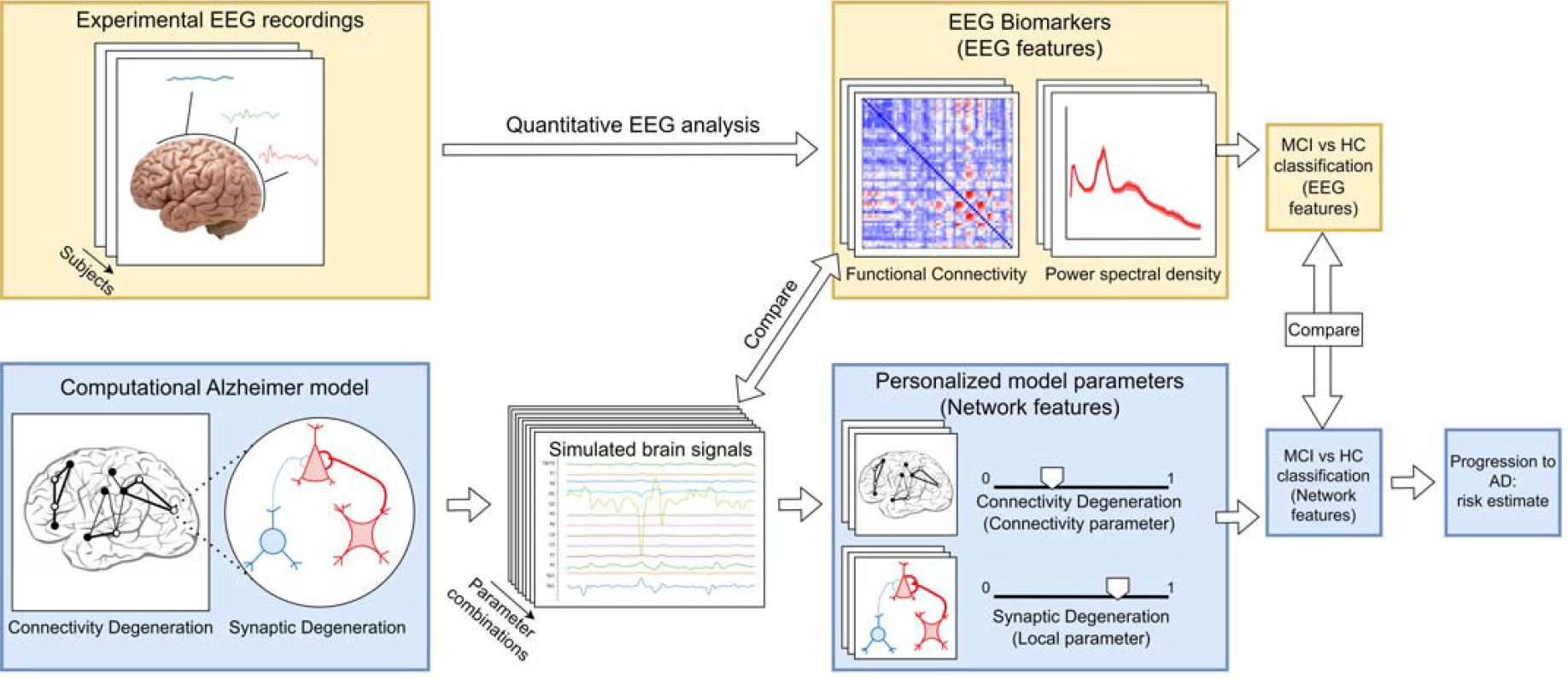
Workflow summary. **Top row**. From left to right: EEG data are acquired from Healthy Condition (HC) subjects and patients with Subjective Cognitive Decline and Mild Cognitive Impairment (MCI). The most group-informative EEG features based on spectral and functional connectivity analysis are extracted and then used to discriminate between HC and MCI. Finally, the same algorithm partitions patients with SCD according to their probability of progressing to AD, deduced from biological hallmarks of the disease (see Methods). **Bottom row**. From left to right: Development of a network model of neurodegeneration in The Virtual Brain frame (see Methods) based on two parameters: a local synaptic parameter and a global connectivity parameter. The values of these parameters for each subject are determined based on the features extracted from their EEG recordings. Discrimination between HC and MCI group is performed in the network features’ space. Partitioning of SCD group is performed in the network features’ space.

## Methods

### Subject recruiting and dementia diagnosis

Subjects included in this study were recruited as part of a clinical neuropsychological-genetic inquiry on pre-AD conditions such as SCD and MCI (ClinicalTrial.gov identifier: NCT05569083). All subjects self-referred to the Centre for Alzheimer’s disease and Adult Cognitive Disorders of the Careggi Hospital in Florence. Inclusion and exclusion criteria follow standard clinical guidelines and are described in detail in the Supplemental Materials. All subjects underwent an exhaustive neuropsychological screening to determine the magnitude of their cognitive complaints. Based on the results of these analyses we divided the initial sample into two groups, with n = 58 subjects classified as SCD (i.e., presence of a subjective, yet persistent deterioration in cognitive capacities with no evidence of such lamentations on clinical cognitive tests) [3]. The other portion of the sample (n = 44) was instead classified as MCI, following the NIA-AA criteria for MCI diagnosis [4]. As a control group, we included n = 17 healthy, age-matched subjects that volunteered to enroll in the study. Local ethics committee approved the protocol of the study, and we collected written consent from all subjects. All procedures related to living human subjects experimentation were done in accordance with both specific national laws and the ethical standards of the Committee on Human Experimentation of the institution, in accordance with the Helsinki Declaration of 1975.

### Candidate markers of disease progression in EEG

EEG recordings and pre-processing are discussed in another work from our group [13].

The candidate biomarkers for disease progression were extracted from two main features of EEG recordings at rest: functional connectivity and power spectral density. Functional connectivity was estimated by computing envelope correlation [21] between electrodes. Envelope correlation is calculated as the Pearson correlation between couples of orthogonalized electrode signals. Briefly, the common phase between the two time series is firstly computed by Hilbert transforming the signals and is then subtracted, thus avoiding the spurious interactions due to the poor spatial resolution of electrophysiological recordings [22]. We computed envelope correlation for delta, theta, alpha and delta bands, and for the whole 0.5-45 Hz range, to study the change in significant broadband connections [23, 24, 25]. FC values were tested for significance and only values larger than half of the average value of FC over all subjects were kept [26]. The candidate biomarkers extracted for each patient from FC were: (i) mean and (ii) standard deviation of FC over all electrode pairs, (iii) count of relevant connections, and average connectivity in the (iv) delta, (v) theta, (vi) alpha and (vii) delta range. PSD was estimated by a Fast Fourier Transform algorithm, using Welch method with Hannning windowing [27] and non-overlapping segments, a resolution of 0.5 Hz was used in both cases. Coherently with previous literature on cognitive decline [11, 12, 22, 28], the candidate biomarkers extracted from PSD were: alpha/delta ratio, alpha/theta ratio, A/LF ratio, individual alpha frequency (IAF). Beta and gamma values were excluded because PSD values in these ranges are orders of magnitude smaller and more disperse than in the lower frequency bands.

### Markers selection

We selected then the EEG features carrying the larger amount of information about the three groups. We computed the mutual information for each feature *f* about the group *c* according to the equations:

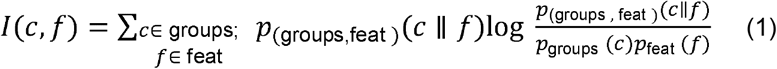

Where *c* is the group, and *f* one of the features of the set *feat*. The two most informative EEG features, one computed from the FC and one from the PSD of the recorded signals (see above) were then selected for classification. See Supplementary Methods for further information.

### Classification algorithm

We developed machine learning algorithms to classify the condition of the patients in the feature space identified by each of the four possible couples of the selected features (see previous subsections and [29]). The classifications were conducted in a supervised fashion, and we trained the algorithm to classify HC and MCI subjects based on the most informative EEG features identified beforehand. Hyperparameters were optimized by means of nested cross-validation. Performances in discriminating HC and MCI were evaluated in terms of both accuracy and F1-score [29]. Outliers, defined as subjects whose computed features were more than four standard deviations far from the mean value, were discarded from both the statistical analysis of the dataset and from the classifying pipeline. After the outlier removal phase, the three groups for the experimental EEG-based analysis remained the same, as we found none, while in the network-based analysis, a single outlier was found in the HC group, that has thus been reduced to n=16 subjects. In a second set of analysis, we used the Random Forest algorithm selected to discriminate between HC and MCI to classify SCD subjects in an unsupervised fashion, using MCI and HC subjects as training set. In this way SCD subjects were partitioned into those closer to HC (SCD->HC) or closer to MCI (SCD->MCI) according to their EEG or network features. Our hypothesis was that the most predictive algorithm is the one in which this partition carries information about the progression of the pathology, i.e., in which SCD->HC have a less severe state than SCD->MCI. Validation of this hypothesis was performed using as severity measure CSF markers (see next subsection). We computed the performance scores with relative confidence values by bootstrapping 100 times the experimental distribution, deriving the range with 95% confidence level (p < 0.05) with Clopper-Pearson method.

### Cerebrospinal fluid analysis

A subset of 23 SCD subjects underwent a lumbar puncture to test for the presence of CSF biomarkers such as *A* β42, *A* β42/*A*β40 ratio, t-tau and p-tau, with Cut-off values determined from the Fujirebio guidelines [30] Patients were rated according to the A/T(N) system [31] and divided in carriers of AD biomarkers (CSF+) and noncarriers (CSF-). We compared then the percentage of carriers (CSF+ subjects) in the two subgroups of SCD: SCD->MCI and SCD->HC (see previous subsection), according to both the EEG-based algorithm and the network-features based algorithm. A significant difference in the ratio of CSF+ subjects between the two sub-populations indicates that the algorithm can classify SCD subjects in terms of perspective of AD progression markers.

### Cortical Model

Healthy cortex was modelled in the TVB framework. Said cortex was modelled as a network of 76 interacting regions. The mesoscopic activity of the region was modelled with the Jansen-Rit neural mass model [32]. Jansen-Rit model depicts the voltage fluctuations due to neuronal activity in cortical structures with a mean-field approach. Cortical regions are depicted as an ensemble of three subpopulations each: one population is of pyramidal cells (excitatory), one is of stellate cells (excitatory) and the last is of interneurons (inhibitory). Mathematically, the activity of these subpopulations, for each node, is given by the solutions of a set of stochastic differential equations. The so-obtained activity is then injected as input in the equations of other regions, connected to the source region by the rules specified by the structural connectivity matrix. The equations of the model are:

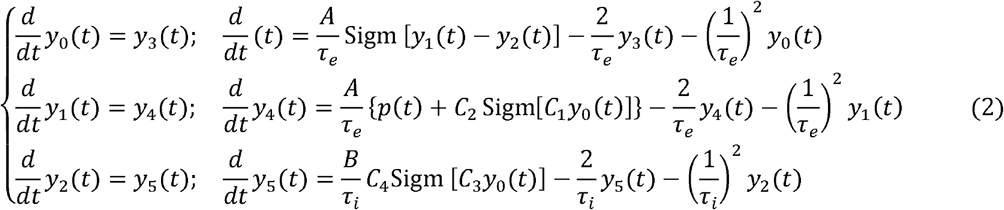

Where *y*_0_, *y*_1_ and *y*_2_ are the post-synaptic potentials of respectively the pyramidal, stellate and interneuron populations, and *y*_3_, *y*_4_ and *y*_5_ are their derivative. *p*(*t*) is the input, written as a firing rate, that is the combination of the activity of other regions *μ*(*t*) and a stochastic term *η*(*t*) that models the physiological noise: *p*(*t*) = η(*t*) + *μ*(*t*).

The Sigmoid term represents the physiology-inspired nonlinear gain function that transforms the average population post-synaptic potential into a mean firing rate:

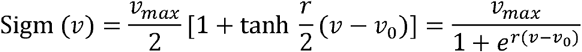

Where *Vmax* is the maximum firing rate of the population, *V*0 is the value of the potential for which there is a 50% firing rate and *r* is the sigmoid slope at *V*_0_. *V*_0_ is also referred to as the excitability parameter of the population. We defined a model to describe the dynamics of each single node of the network, and then we linked these nodes by using a structural connectivity matrix, in which each couple of regions is characterized by a connective weight *C*_*weight*_ that mathematically transcribes the,number of axonal fibers connecting the regions, and a distance between regions called *C*_*lenght*_ that describes the length of the fibers. Both the model and the connectivity matrix we utilized from the TVB data-frame (33). In particular, the structural connectivity matrix and the cortex parcellation are derived from the standard TVB atlas, which combines high precision and reliability [33]. The of weights *C*_*weight*_ the TVB connectome are assigned integer values ranging from 0 to 3, that transcribe null (*C*_*weight*_ =0), weak (*C*_*weight*_ = 1), medium (*C*_*weight*_ = 2), and strong (*C*_*weight*_ = 3) anatomical connections. The connectivity constants *C*_1,2,3,4_ are proportional to the number of synapses that link the sub-populations together. The constants *A* and *B*, expressed in mV, are the maximal amplitude of the post-synaptic potentials for excitatory and inhibitory neurons while, *τ*_*e*_, *τ*_*i*_ and expressed in ms, are the time constants of excitatory and inhibitory population, which τ τ lump together dynamic features such as delays of the synaptic transmission, and the delay in signal transmission due to the propagations along the dendritic tree. These last two parameters are those that we altered in order to depict the progression of the disease, and also the major reason why we chose this model, due to the fact that tuning the τ_*e*_ τ_*i*_ implementing the excitation/inhibition imbalance *τ*_*e*_/*τ*_*i*_ ratio is a simple yet highly effective way of in AD [34]. Model parameters are recurrent summarized in Table 1. Functional connectivity was computed for simulated EEG as Pearson correlation between brain regions’ activity. Power spectral density across frequency bands was obtained by Welch method as we did for experimental signals.

**Table 1:**
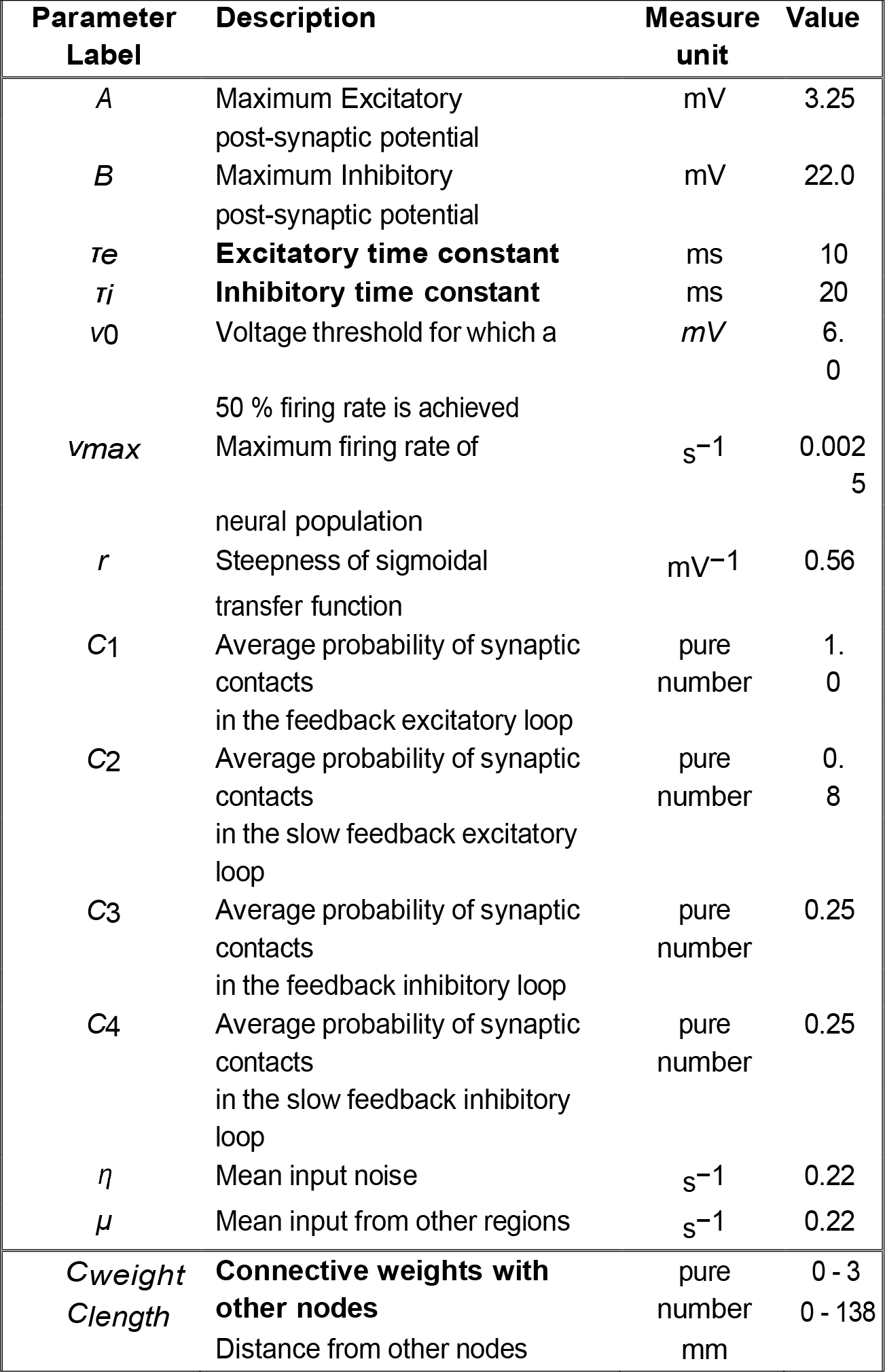
Model parameters. The bold-highlighted quantities are those modified in order to depict AD effects (by using the network features used to determine condition severity, see main text).

### Disease progression model

We modeled the progression of pathology with two quantities: a local parameter (*lp*) of regional degeneration, and a connectivity parameter (*cp*) of connectome degeneration. These quantities describe respectively the alteration of white matter fiber, in terms of both white matter atrophy and neuroplasticity [35, 36] and the degeneration of synaptic transmission [20] according to the equations:

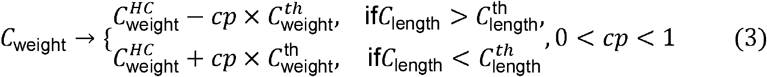

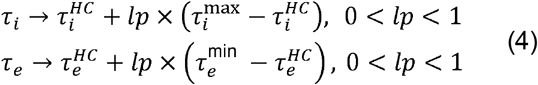

Where 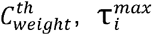 and 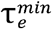 are the extremal values of the highlighted quantities, and are 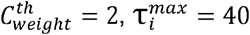, ms, 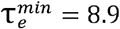 ms, deduced from biophysical constraints [32, 33]. Note that we introduced, alongside excitation/inhibition imbalance and white matter fibers atrophy, a model of the correcting mechanisms operated by the brain in order to cope with the disease, such as synaptogenesis and neural plasticity [36, 37, 38]. This effect is modelled as an increase in short range connection (i.e an increase in connective weights of the structural connectivity matrix) with initially impaired regions, that is introduced along with the reduction in long range connections implemented for the same regions (Eq. 3). The discrimination between short and long connections is made on the length of axonal tracts connecting the regions, *C*_*length*_ with short (long) connections existing between regions that are separated by a distance smaller (greater) than a threshold, that we deduced to be equal to 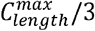 from anatomical considerations of the utilized connectome [33]. AD appearance is not ubiquitous in the brain, and there is a subset of cortical regions in which the disease spreads significantly faster than in others [39]. To model this spreading anisotropy we induced the connectome and *τi* alterations only in a subset of regions, mostly temporal regions taken from well-known Braak stages, while τ*e* modifications appear in the whole cortex (Eq. 4).

These initially impaired regions are orbitofrontal and polar regions of prefrontal cortex, and the polar, central, inferior, and ventral temporal cortex.

### Brain signal simulation

Regional activity is obtained by solving the set of stochastic differential equations given by the Jansen-Rit model (Eq. 2), which are computed by means of the stochastic Heun method [40], with a timestep of 0.1 ms. The stochastic term η(*t*) is an additive, Gaussian white noise equal for all regions. For each run, we simulated 10 s of local electrophysiological activity, with a resolution of a ms.

### Tuning the model to single subject data

We estimated local and connectivity parameters for each individual patient as follows. First, we computed in simulated data A/LF ratio and average BFC as a function of *lp* and *cp*. We did a grid-search by changing both the connectivity and local parameters *lp* and *cp* in 50 linear steps between 0 and 1. We thus obtained the two matrices *A/LF^mod^*(*lp, cp*) and *BFC*^*mod*^(*lp, cp*).

We then normalized both the experimental and the simulated data between their minimum and maximum values, we computed the total weighted euclidean distance between the experimental values of the EEG features of each subject *sub*: *A/LF^exp^* (sub) and *BFC*^*exp*^ (sub) and each point of the simulated matrices:

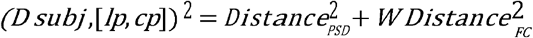

Where the quantities *Distance*_*PSD*_ and *Distance*_*FC*_ are defined by the equations:

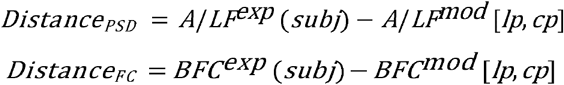

The purpose of *W* is to find the weighted distance from the experimental EEG features, that, when minimized, identifies the distribution of network features that best classifies HC and MCI subjects. Each value of *W* univocally identifies a distribution of *cp* and *lp*. We determined the optimal *W* by looking for the distribution that maximized the HC-MCI classification performance in the *W* value range 0.005 - 16. We found *W* = 12 to be the best weight value, and we then determined the *lp*-*cp* combination for each patient, by minimizing the distance with that value of *W* as weight. By identifying this values’ pairs, we unequivocally find the combination of network features that best describes the given patient in term of the relevant features computed from their EEG. We also checked the robustness of this procedure, by applying a gaussian random noise to the experimental *A/LF^exp^* (*sub*) and *BFC*^*exp*^ (*sub*) values used to determine the best fitting parameters. The noise was distributed for each feature in an interval centered in its original value, with the interval length given by 10 % of the same value. We then recomputed the network features from the new distribution of EEG features.

We hereby report for the sake of clarity the pseudocode of the function used to determine the best fitting parameters for each subject:

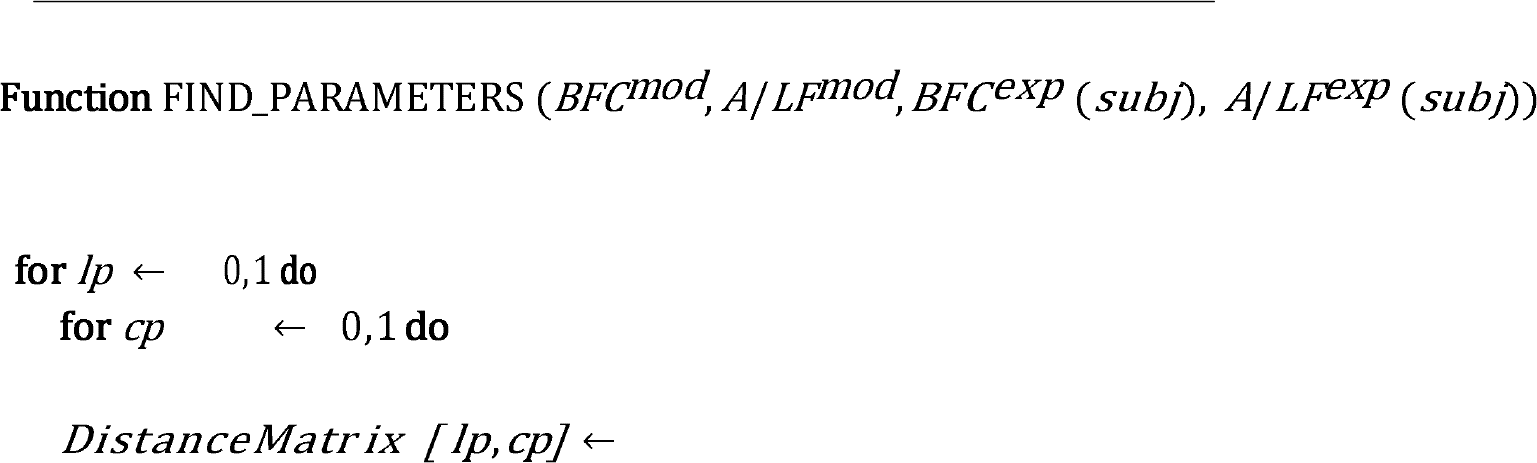

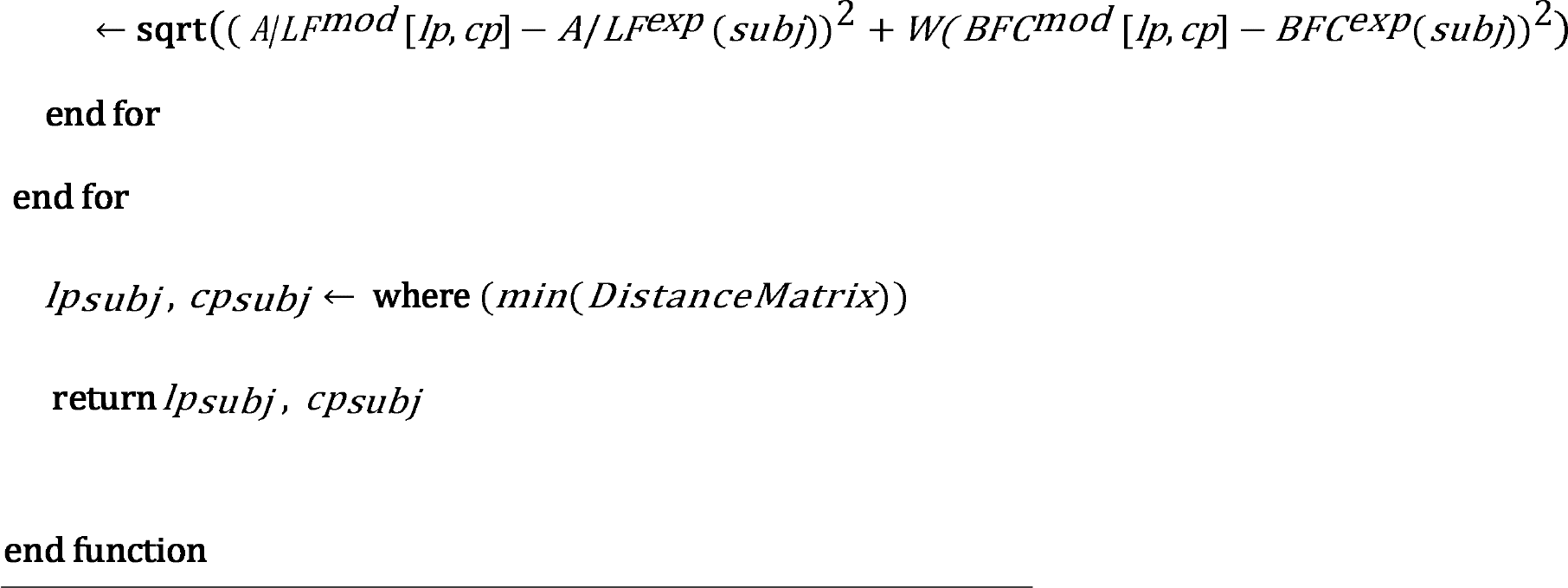

### Classifying patient conditions with network parameters

The classification of the patient conditions described in the subsection “Classification Algorithm” was then repeated based on the network parameters *lp* and *cp* rather than the EEG features *A/LF^exp^* and average alpha FC. First, the patients’ specific values of *lp* and *cp* were used to discriminate between HC and MCI. Then, SCD patients were assigned by the same algorithm either to the HC or to the MCI category as we did for EEG features. Finally, to check if our model could determine the degree of structural alterations due to disease progression, we compared the percentage of CSF+ subjects in the two subsets identified by the classifier (see Cerebrospinal fluid extraction). This was done to assess if our proposed classification and prediction could outperform the one based on standard experimental data.

### Graph theoretical analysis

To link the functional and dynamic non-linearities observed during the simulated structural connectivity degeneration, we performed a graph-theoretical analysis of the structural matrices of the model while varying the *cp* parameter. We utilized the Randic index [41], a quantity that has already been used in studies concerning AD [42].

This quantity, defined as the sum of 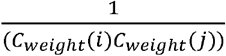 out of every possible (*i,j*) pair is anti-correlated with the tendency of each node to be linked with nodes of similar degree, the so called “Rich-club phenomenon” [43]. The Rich-club phenomenon is known to apply to the connections in the healthy cortical network [43]. We studied the evolution of Randic index during the progression of the disease, by computing this quantity for every value of *cp*, comparing the results with the variation of both *A/LFmod* and *BFCmod* according to the same variable. We also investigated the Randic evolution during disease progression in a network model with random structural connectivity matrix, to assess the differences with the physiological case and check if the non-linearities observed experimentally could be derived from the non-trivial topology of the network.

### Statistical analysis

Distributions normality was assessed by using Shapiro-Wilk test. For approximately normally distributed variables (p > 0.01) we used one-way ANOVA f-test, otherwise we chose the Kruskal-Wallis non-parametric test to compare the three groups. Similarity in distribution of connective weights was assessed with Kolmogorov-Smirnov test. Post-hoc analysis was conducted by Bonferroni correction after pairwise comparisons between groups. All tests were two-tailed, except for the test on the significance of the CSF+ prediction, that was a one-tailed test on the binomial distribution with 50% probability as null hypothesis, and the significance test for the performance metrics of machine learning pipeline. The confidence ranges of performance metrics for MCI vs HC classification were assessed by generating a null distribution with the bootstrap test with 250 iterations and by taking the 2.5% and 97.5% percentiles (p = 0.05). For the confidence ranges of CSF prediction, since the sample sizes were smaller than in the previous case, we opted for Clopper-Pearson method. Statistical analysis has been implemented in Python by using the standard *Scipy* package.

## Results

### EEG features in early stages of dementia

Forty-four subjects with Mild Cognitive Impairment (MCI), fifty-eight with Subjective Cognitive Decline (SCD) and seventeen Healthy controls (HC) performed EEG recordings for at least 30 minutes at rest (see Methods for details). We first classified patients in these groups based on EEG features, extracting for each patient the power spectral density (PSD) and the FC between pairs of electrodes, two known EEG biomarkers of AD [11, 12].

The PSD significantly differed across the three groups (see Methods) only within the alpha band ([8-12] Hz) and within the low-beta range ([12-20] Hz) (p<0.05, one-way ANOVA with Bonferroni correction, Fig. 2A). In both ranges the PSD was higher for HC than for SCD and for SCD than for MCI. The opposite ranking was observed for delta ([0.5-4] Hz) and theta ([4-8] Hz) bands although inter-group differences were not significant. We computed the information about the three groups carried by other spectral EEG biomarkers based on standard frequency bands (see Methods), and we found the alpha/(delta+theta) power ratio (A/LF ratio) to be the most informative EEG spectral feature (see Supplementary Fig. 1a). However, this ratio was not significantly different across groups (Kruskal-Wallis test, statistic = 2.86, p=0.24, Fig. 2B). We computed FC (see Methods) over frequency bands, along with broadband FC [23,24,25] (BFC, see Supplementary Fig. 2). Mean BFC values did not differ significantly between groups (0.072 ± 0.037 for the HC group, 0.071 ± 0.035 for the SCD group and 0.069 ± 0.031 for the MCI group). We then binarized the BFC setting as threshold the mean BFC value [26], to study the differences in the number of relevant functional connections between groups (see Methods). MCI group displayed an average decrease of significant connections compared to HC (17.8% of connections present in HC are lost in MCI while only 7.6% of the connections present in the MCI group are gained in the HC one, see Supplementary Fig. 3a left). Surprisingly, comparing SCD to HC, the overall prevalence was an increase in the number of significant connections (11.9% gained vs 6.9% lost, Supplementary Fig. 3a center). Connections were instead lost in the MCI group compared with the SCI group (18.2% lost vs 2.95% gained, Supplementary Fig. 3a right). This suggests a non-monotonous progression of functional connectivity weakening across dementia severity [36, 37, 38]. We computed then the amount of information carried about the three groups of the EEG functional connectivity biomarkers of AD (Supplementary Fig. 1b). The most informative was the average alpha band FC even if no significant difference was present between groups (Fig. 2C, Kruskal-Wallis test, statistic = 5.70, p = 0.057). The shape of the BFC distribution, instead, significantly changed across groups (Kolmogorov-Smirnov test, HC vs SCD: test = 0.077, HC vs MCI: test = 0.10, SCD vs MCI: test = 0.038, p << 0.0001 across all groups), with the largest variability being present in SCD subjects (Coefficient of variation was 0.051 for SCD, while 0.047 for both HC and MCI). We tested the classification performance of combinations of PSD and FC features in discriminating between HC and MCI (Table 2) using Random Forest as it outperformed other approaches (Supplementary Table 1). The strongest classification was based on average alpha band FC and A/LF ratio (see scatter plot in Fig. 2D), achieving an accuracy of 78% [69%-86%] (alpha=0.05 computed from bootstrap distribution, see Methods), with f1-score of 0.85 [0.76-0.91] and recall (fraction of MCI subjects correctly identified) of 0.82 [0.73-0.91]. The HC-MCI inter-cluster separation of 0.46 (see Methods). We investigated whether we could discriminate SCD subjects more at risk of progressing toward dementia based on their similarity with MCI rather than with HC in the EEG features space. We used then the same algorithm used to discriminate between HC and MCI (using again HC and MCI groups as training set) to classify SCD subjects in an unsupervised fashion (see Methods). In this way SCD subjects were partitioned into a subset classified as being closer to the healthy condition (SCD->HC) and a subset closer to MCI condition (SCD->MCI). We hypothesized that this classification reflected the outlook of SCD patients. We tested this hypothesis by comparing the results of cerebrospinal fluid (CSF) markers (strongly related to structural alteration [7], see Methods) for the two subgroups. However, the percentage of CSF+ was almost the same (33% in SCD->MCI subgroup and 36% in the SCD->HC subgroup). The accuracy of the algorithm in discriminating CSF+ and CSF-was 48% [38%-58%] (Clopper-Pearson method, see Methods), with f1-score of 0.40 [0.30-0.50], and recall of (fraction of CSF+ subjects correctly identified) 0.50 [0.39-0.61] (see Methods). Finally, at 1y follow-up, three of the SCD subjects transitioned to MCI. Only one of them belonged to the (SCD->MCI) set. This suggests that decoders based on standard PSD and FC EEG features display a low segregation between MCI and HC and could not subserve an outlook of SCD subjects condition.

**Table 2:** Top-4 classifier performances trained with combinations of EEG spectral and FC feature.

| EEG features | Classifier | Accuracy | F1-score |
| --- | --- | --- | --- |
| **A/LF ratio - FC alpha band average** | Random Forest | 0.78 | 0.85 |
| A/LF ratio - FC alpha band average | SVM, rbf kernel | 0.68 | 0. |
| A/LF ratio - BFC average | SVM, polynomial | 0.67 | 0.71 |
| Alpha/theta ratio - BFC average | SVM, linear | 0.63 | 0.65 |

**Fig. 2:**
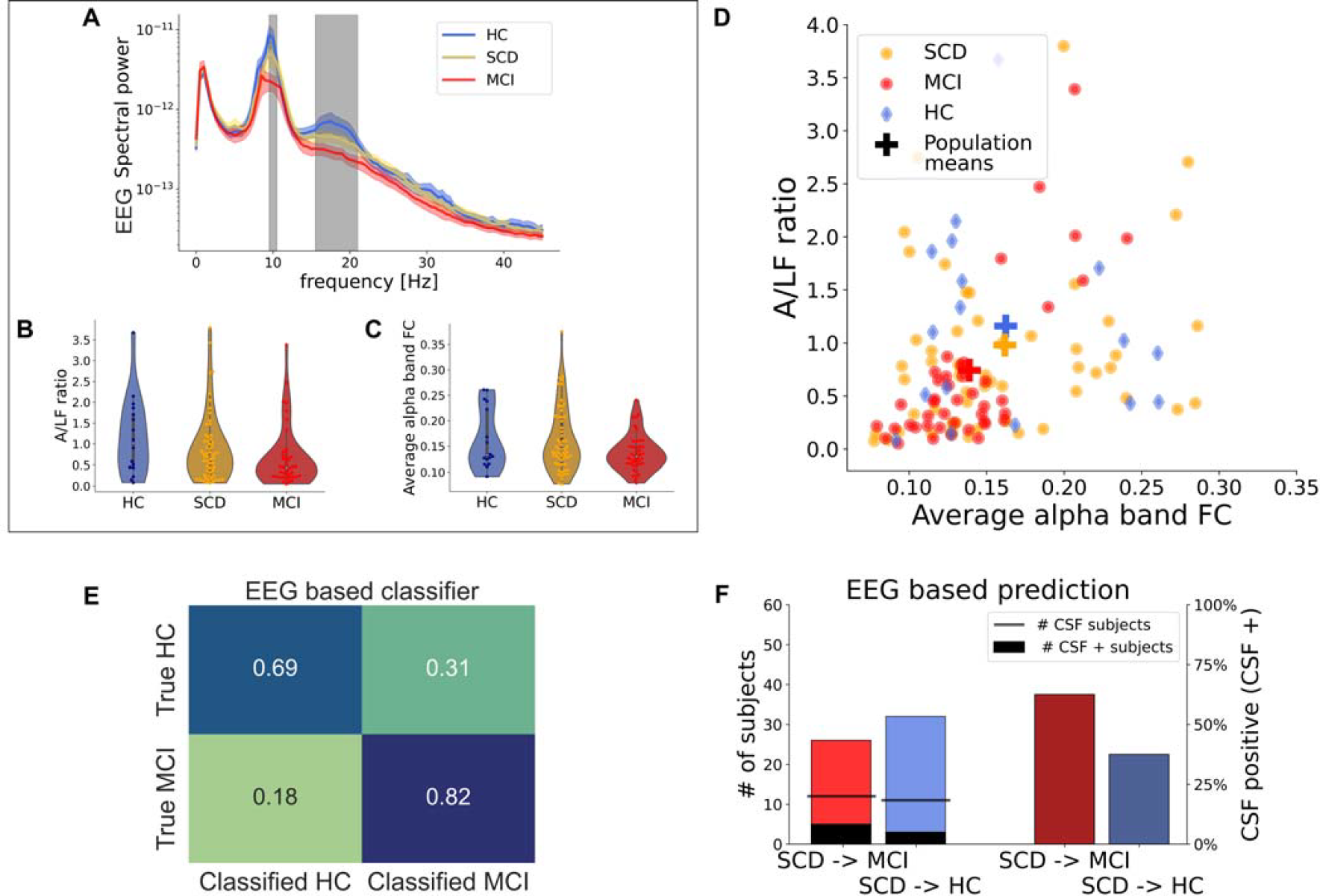
Decoding of patients’ condition and prediction based on EEG features. **(A)**: Mean PSD for all groups, shaded area around PSD curves represents the standard error of the mean. Gray bars indicate frequency ranges displaying significant differences (p <0.05) across groups. **(B)**: Ratio between power in the alpha band and lower frequencies (A/LF ratio) for the three groups. **(C)**: Functional Connectivity in the alpha band (alpha FC) for the three groups. **(D)**: Combined representation of A/LF ratio and average alpha FC for each subject in each group. Cross-shaped markers indicate cluster centroids for the three groups (see Methods). **(E)**: Confusion matrix for HC vs MCI for optimal classification algorithm (see Table 2) based on EEG features **(F)**: Left: SCD subjects classified as HC (light red) and MCI (cyan) with the same classification algorithm used in (D) (see text). Black bars indicate the numbers of subjects whose CSF was positive to AD-biomarkers (CSF+) within each set. Right: percentages of CSF+ subjects for each category (brown red and dark cyan).

### Multiscale cortical network model of progression to dementia

To overcome the limitations of EEG in assessing dementia severity, we hypothesized that reconstructing from the model the specific circuital changes induced by the early stage of AD-type dementia in each subject (Fig. 1, bottom) could highlight parameters enabling a more accurate classification of SCD patients than the one allowed by EEG features. We started from the model of the healthy cortex available in the EBRAINS The Virtual Brain platform (TVB, see Methods). Then we modeled the neural circuit correlates of the onset of AD-type dementia considering changes at the level of both local dynamics and global connectivity (Fig. 3 and Methods). Briefly, structural connectivity degeneration was modelled by considering both a decrease in long-range connections due to white matter atrophy and an increase in short-range connections due to neuroplastic mechanisms (see Methods). Synaptic degeneration was accounted for by modelling alterations in inhibitory and excitatory synaptic time scales. These two processes are summarized by two network features: the connectivity parameter *cp* for long-range effects and the local parameter *lp* for short-range effects (see Eqs. 3 and 4 in the Methods and in Fig. 3).

**Fig. 3:**
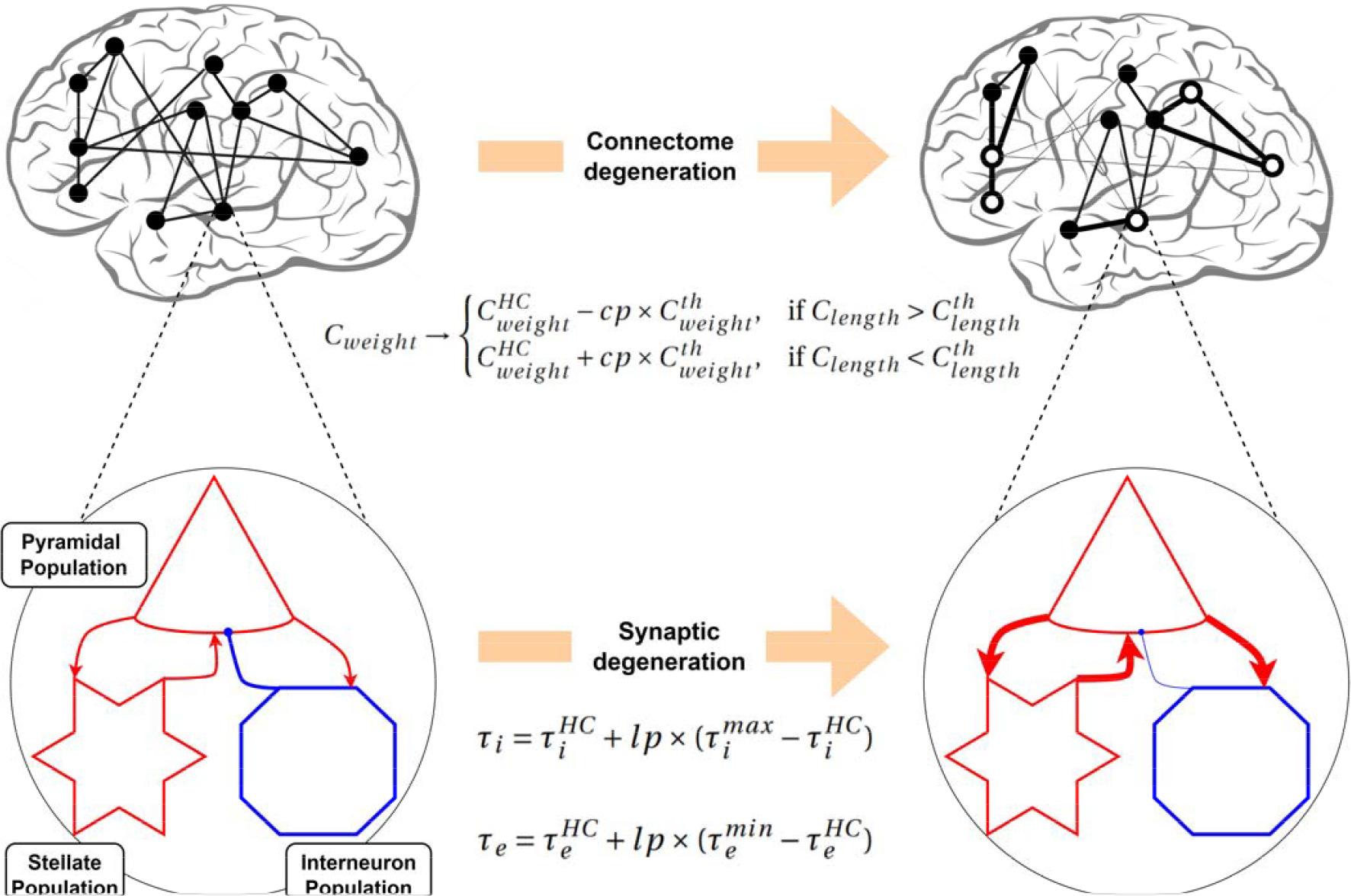
Outline of the model of dementia-related structural alterations. **Top row**: A macro-scale connectivity parameter (*cp*) describes the degree of alterations in structural connectivity, through linear strengthening of short range and weakening of long-range connections (see Methods). Thickness of lines is proportional to connection strength; white nodes are those affected by AD-type dementia. **Bottom row**: A meso-scale local parameter (*lp*) describes the degree of synaptic degeneration through a local slowing of inhibitory synapses, alongside a global quickening of excitatory synapses. This leads to hypo-inhibition in the regions impaired by the disease, along with a milder but ubiquitous hyperexcitation.

We first assessed whether the model was able to reproduce the alterations of EEG features (PSD and FC) observed in SCD and MCI groups. For suited pairs of *cp* and *lp* values (see Methods) the model was able to reproduce qualitatively both connectivity and spectral features of HC, SCD and MCI from EEG recordings. The combination of cp and lp parameters reproducing experimental features was [*cp*_*HC*_ = 0.48, *lp*_*HC*_ = 0.30] for HC, [*cp*_*SCD*_ = 0.64, *lp*_*SCD*_ = 0.46] for SCD and [*cp*_*MCI*_ = 0.98, *lp*_*MCI*_ = 0.84] for MCI, coherently with the expected positive correlation between the magnitude of network features and the severity of the condition. For these network features pairs the model displayed the experimentally observed decrease in alpha band in SCD and MCI groups with respect to the HC group, as well as the decrease in beta band power observed in MCI subjects (12.5% ± 2.6 power loss with respect to HC values for the MCI group in the beta band, respectively 10.3%±2.1 and 41.3%±8.3 power loss for SCD and MCI with respect to HC values in the alpha band). Coherently with the experimental results, A/LF ratio decreased for larger values of network features (Supplementary Fig. 3a).

Since in the network model we computed FC between cortical regions rather than between electrodes, we compared the simulated network FC with the broadband envelope correlation computed from EEG signals, as previous studies suggested that this measure is suited to study functional connections between source regions [44]. Note that BFC was a highly informative EEG feature discriminating the groups (Table 2, Supplementary Fig. 1). For the same parameter values reproducing observed spectral modulations the model also displayed the same observed differences in FC across groups (SCD vs HC: 8.3% lost vs 12.1% gained connections, MCI vs SCD: 15.5% lost vs 6.95% gained connections, MCI vs HC: 13.1% lost vs 8.30% gained connections, Supplementary Fig. 3b-c). Coherently with the experimental recordings analysis, average broadband functional connectivity displayed similar values across groups (0.074 for HC and SCD and 0.071 for MCI). Overall, the network model was able to capture both variation in the low frequency spectrum and the non-linearity in connectivity progression associated with different conditions in EEG recordings.

### Relationship between dementia-related network features and dementia-related EEG features

We investigated then the relationship between the EEG features of average BFC and A/LF ratio in simulated data and the underlying network features modeling the degeneration process *cp* and *lp* (Fig. 4). We simulated the EEG as a function of both *lp* and *cp*, and we computed the associated BFC and A/LF ratio.

**Fig. 4:**
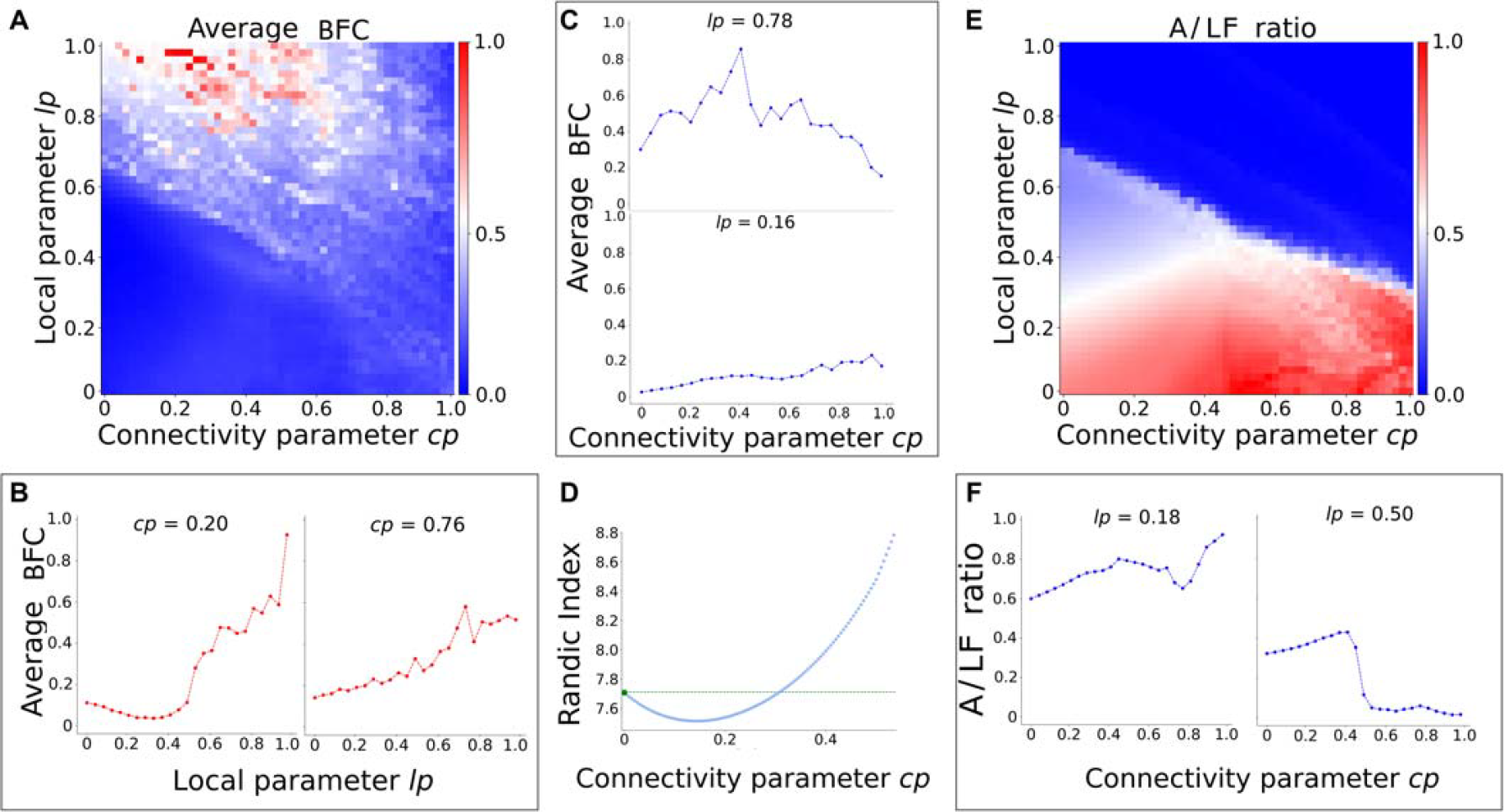
Relation between network features and EEG features. **(A)**: Average broad-band functional connectivity (BFC) for the network model as a function of local and connectivity parameter, *lp* and *cp*. **(B)**: BFC as a function of *lp* for fixed values of *cp*. **(C)**: BFC as a function of *cp* for fixed values of *lp*. **(D)**: Randic index as a function of *cp*, showing nonlinear evolution of network assortativity during disease progression (see Methods). **(E)**: A/LF power ratio for the network model as a function of *lp* and *cp*. **(F)**: A/LF ratio as a function of *cp* for a fixed value of *lp*.

Broadband Functional Connectivity (Fig. 4A) increased with *lp* for all values of *cp* (Fig. 4B). The relationship between BFC and *cp* depended instead on *lp*: when *lp* was high BFC displayed a peak value for intermediate values of *cp*, suggesting a non-linearity of BFC during progression toward dementia, while when *lp* was low BFC increased monotonously (Fig. 4C). To understand how this was related to the structure of the network we measured the assortativity of the network computing its Randic index (see Methods and [41,42]) as a function of *cp* (Fig. 4D). The Randic index displayed a non-linear trend, with values even lower than those found in the healthy case for early stages of the disease progression, followed by a steep increase for high valued of *cp* increases. The Randic index links the state of structural and functional alterations and could explain the non-linearity observed in the FC measurements in EEG. Our model suggests that the gradual decline of long-range axonal fibers and the enhancement of connections between nearby affected regions could cause a temporary increase in assortativity. This is then followed by a sharp decrease due to the worsening state of long-range connections. Our findings suggest that the topology of a healthy cortical network is the source of observed non-linearities indicated by the Randic index. We tested this hypothesis by progressively increasing the cp parameter, this time starting from a random structural connectivity matrix, and we found both the average functional connectivity and Randic index to show monotonic evolution with the progression of the disease, suggesting that the nonlinear evolution observed in experimental data stems from the complex topology of the network (Supplementary Fig. 4a-b).

We investigated then the evolution of the A/LF ratio when the parameters associated with dementia progression are increased (Fig. 4E). For high values of *lp* the ratio decreased with *cp*, while the opposite was true for low values of *lp* (Fig. 4F). The increase of *lp* was instead always associated to a steep decline in the ratio.

### Personalized network features lead to reliable classification and prediction of structural alterations

We finally proceeded to compute personalized network features *cp*_*subj*_ and *lp*_*subj*_ to feed them (rather than EEG features) into the classification and prediction algorithms.

We determined for each subject the combination of network parameters [*cp*_*subj*_ *lp*_*subj*_] for which the features of the simulated electrophysiological signals were closer to the actual EEG features computed from the recordings of the subject. This allowed us to define for each subject a personalized network with an estimated measure of connectome degeneration, given by *cp*_*subj*_, and an estimated level of synaptic degeneration, given by *lp*_*subj*_(Fig. 5A). The two parameters were correlated across subjects for merged groups (r^2^= 0.077, p = 0.002), but correlation decreases when computed for each group (r^2^ = 0.40 p = 0.006 for HC, r^2^ = 0.12, p = 0.008 for SCD and r^2^ = 0.083, p = 0.061 for MCI) with a larger mean squared error (MSE) for SCD and MCI groups (MSE = 0.12 for HC, 0.91 for SCD and 0.62 for MCI see Fig. 5B).

**Fig. 5:**
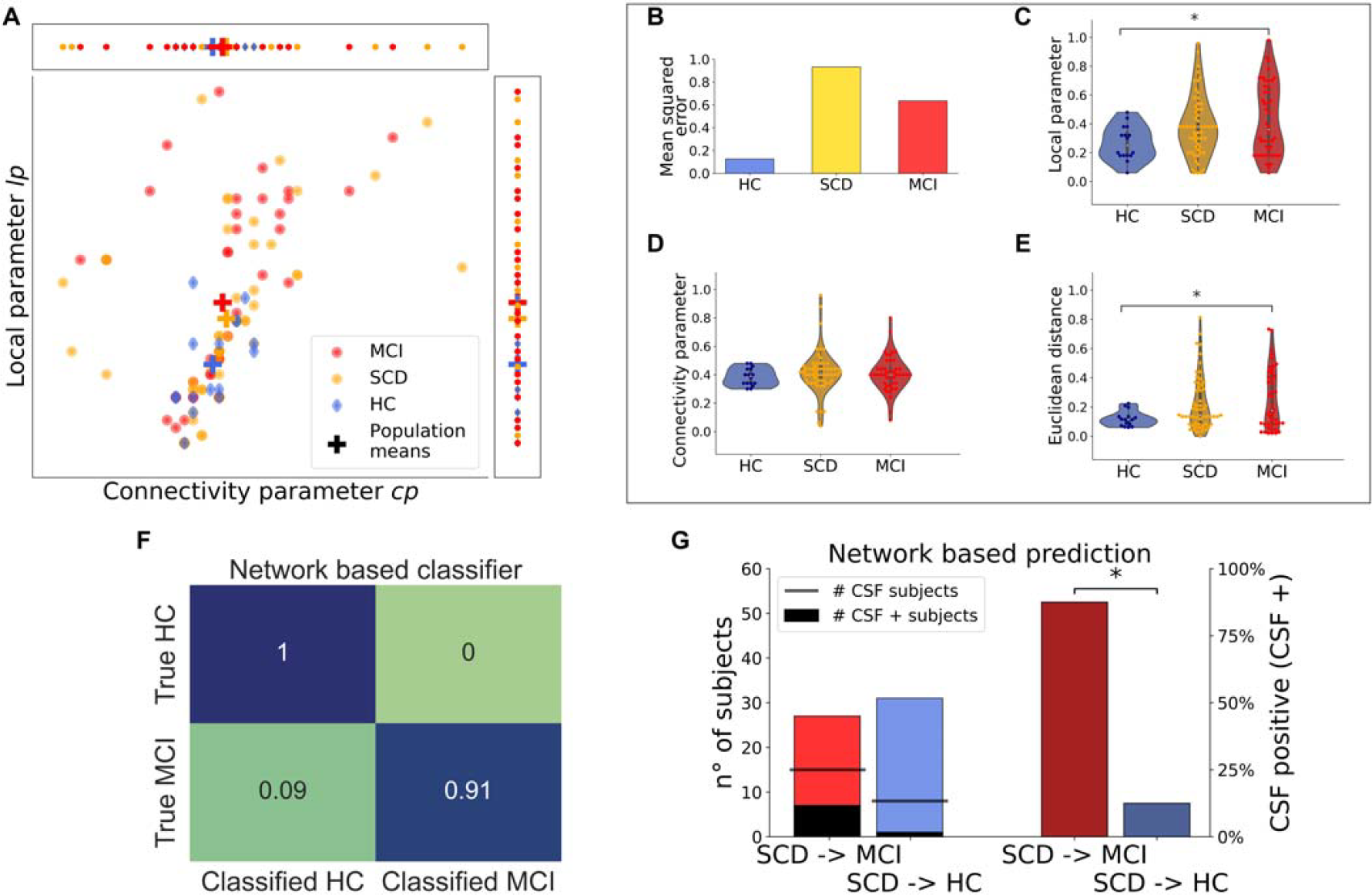
Decoding of patients’ condition and prediction based on network features. **(A)**: Combined representation of network features *lp* and *cp* for each subject in each group. Cross-shaped markers indicate cluster centroids for the three groups. **(B)**: mean squared error of the linear fit for each group (bottom panel). **(C-D-E)**: Distribution across groups of local parameters (C), connectivity parameter distribution (D), and Euclidean distance from the healthy cluster center (E). **(F)**: Confusion matrix for HC vs MCI optimal classification algorithm based on network parameters. **(G)**: Left: SCD subjects classified as HC (light red) and MCI (cyan) with the same algorithm used for experimental EEG features (see text). Black bars indicate the number of CSF+ subjects within each set. Right: percentages of CSF+ subjects for each category (brown red and dark cyan). Significance notation as in Fig. 2.

We found *lp* to be significantly smaller in HC subjects with respect to MCI (one-way ANOVA, f = 6.17, p = 0.048, with Bonferroni correction) but not to SCD (one-way ANOVA, f = 1.73, p = 0.126, with Bonferroni correction) subjects (Fig. 5D). The parameter *cp* did not differ significantly across groups (Kruskal-Wallis test, statistics = 0.35, p = 0.70) (Fig. 5E). The HC group was well separated from the others as the average Euclidean distance in the [*cp*_*subj*_ *lp*_*subj*_] space (Fig. 5A) from the centroid of the HC group (see Methods) was significantly smaller in HC subjects with respect to MCI (one-way ANOVA, f = 6.56, p = 0.039, Bonferroni correction) and to SCD (one-way ANOVA, f = 4.89, p = 0.09, Bonferroni correction) subjects (Fig. 5D). We also tested the robustness of this pipeline by checking how the assigned parameters change after adding a gaussian white noise to initial data (see Methods). Results are shown in Supplementary Fig. 5, that highlights the stability of the parameters determination procedure.

The fact that network features displayed significant differences across groups is particularly relevant as this was not the case with the experimental EEG features. To assess whether this led also to an improvement in the classification performance, we repeated the classification between MCI and HC subjects using the same algorithm utilized for the classification with EEG features (see Methods) but using only the network features extracted subject-wise. We found an accuracy of 93% [87% 97%], with f1-score of 0.95 [0.91 0.98] and a recall (fraction of MCI subjects correctly identified) of 0.91 [0.84 0.96]. The two clusters also displayed a clear separation *d*^*mod*^ = 1.26.

Since the algorithm based on network features performed better than the EEG-based one in discriminating between HC and MCI, we investigated whether it could determine the severity of the condition of SCD subjects and possibly their future progression. Following the procedure applied for the EEG features based algorithm (see above and Methods), we partitioned the SCD subjects into SCD classified as MCI (SCD->MCI) and SCD classified as HC (SCD->HC) based on their personal network features. To test the hypothesis that the network features-based partition could predict the state of structural alterations of SCD subjects, we compared the presence of CSF markers for the two SCD subgroups. The data size was too small to perform a Fisher exact test, but we found that the percentages of CSF biomarker carriers in the SCD->MCI group was almost four times the percentage observed in the SCD->HC classified group (47% vs 12%, p = 0.04, Fig. 5G). The accuracy of the partition in discriminating CSF+ and CSF-was 61% [51% – 70%], the f1-score 0.60 [0.50 0.69], and the recall was 0.87 [0.78 0.93]. These results suggests that an algorithm based on the network features extracted from resting EEG signals could determine the state of structural alterations induced by the disease, both across clinical groups and within them. Strikingly, 3/3 subjects that converted from SCD tom MCI at 1y follow-up belonged to the set for which the network model predicted a negative outcome, while only one was predicted with EEG features. This corroborates the strong enhancement in diagnostic and prognostic ability obtained by the network-driven analysis of EEG signal in comparison with standard quantitative techniques.

Overall, the network-based algorithm significantly outperformed the EEG-based algorithm in HC vs MCI classification in f1 score (Fig. 6, top row), while for the CSF+ prediction in SCD subjects, all performance metrics were significantly greater in the network-based algorithm (Fig. 6, bottom row). Note that this is non-trivial as the network features replaced the EEG features, rather than being used in combination with them to enhance the performance of the classifier (see Discussion).

**Fig. 6:**
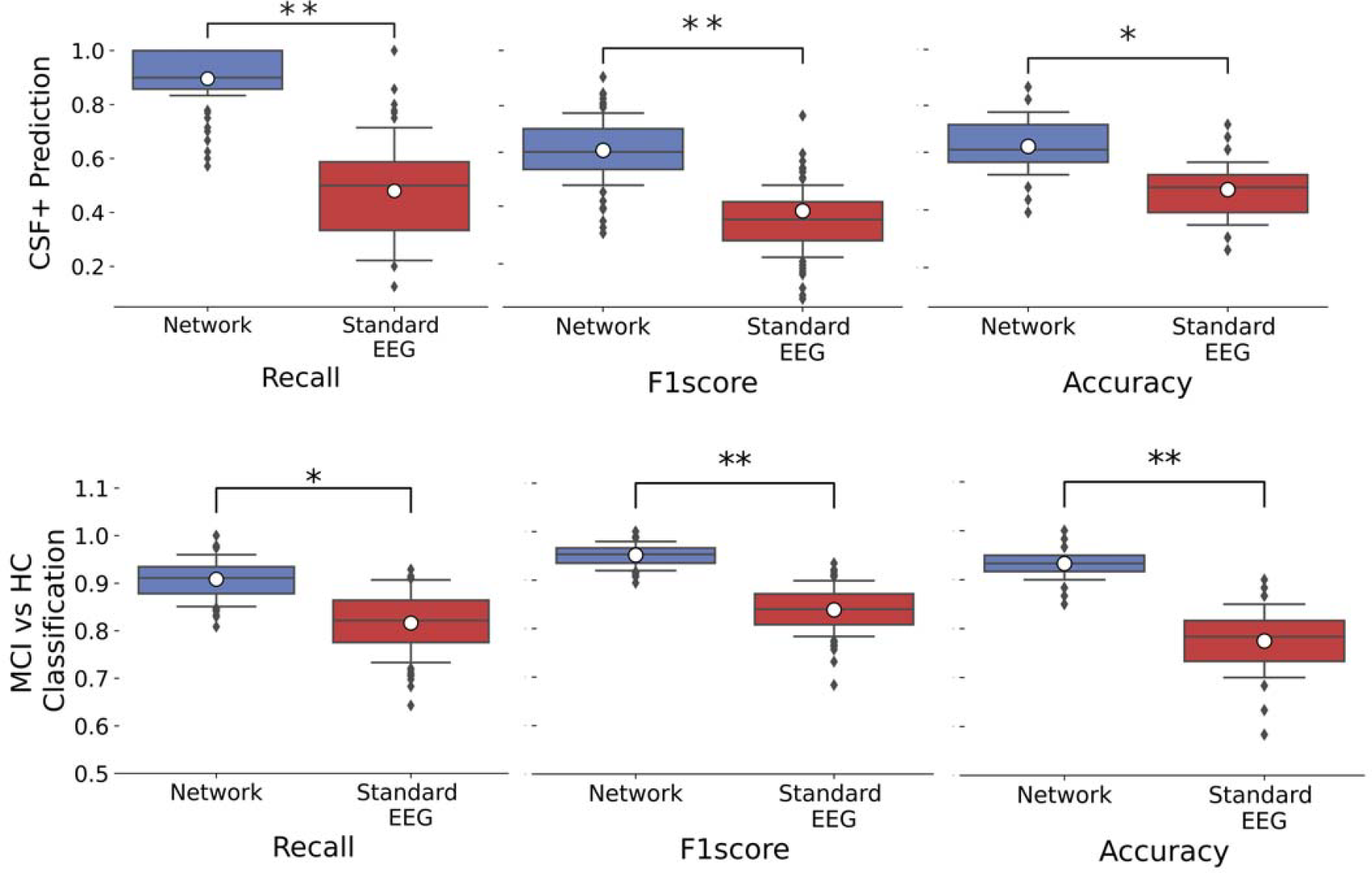
Performance comparison between EEG-based and Network-based algorithms after bootstrapping. **(Top row)**: Performance on predicting the biological hallmarks of the disease in the CSF; **(Bottom row)**: Performance on classifying between MCI and HC conditions. White dots represent experimental values. Significance notation: * assigned when both conditions: (i) experimental value of EEG-based algorithm below confidence range of the network-based algorithm and (ii) experimental value of network-based algorithm above confidence range of the EEG-based algorithm were true (p < 0.05). ** stands for non-overlapping confidence ranges at p = 0.05.

## Discussion

We developed a computational model estimating the level of degeneration in local connections and global connectivity during the trajectory of dementia. These changes can be estimated patient-wise combining the model with spectral and connectivity attributes of individual resting-state EEG recordings. Leveraging these attributes as input, machine learning algorithms demonstrated prowess in classifying early stages of dementia and forecasting their progression, significantly surpassing algorithms founded solely on conventional informative EEG features. The potency of our approach stems from its innovative incorporation of knowledge concerning the course of AD-related neurodegeneration in the decoding of EEG signals.

### A subject-dependent, multi-scale model of disease progression

Our model adeptly replicates salient aspects of healthy and pathological EEG recordings across varying scales, aptly encompassing the spectral and connectivity characteristics observed experimentally. The model captures the dynamic spectrum of anomalies wrought by the disease through modulation of local circuit anomalies encompassing synaptic degeneration and hyperexcitation, alongside the integration of global effects such as white matter atrophy. While an earlier work on AD modeling introduced a specific model pertaining to the decline in the alpha/theta ratio [19], our model extends its scope to accommodate a broader array of structural modifications, thereby becoming capable of accounting for their effects on connectivity. Furthermore, we have introduced explicit representation of neuroplastic adaptive phenomena— embodied as augmented short-range increase in structural connectivity within impaired regions—aligning with the observed nonlinear evolution of functional connectivity as dementia advances. These structural and circuital shifts, distilled into two parameters (cp and lp), readily calibrate the model to each patient’s electrophysiological signals, thereby facilitating linkage between experimentally observed disease hallmarks and computationally determined structural deviations. Distinct from prior work such as [19], which focused on predicting EEG features from structural changes, our endeavor prioritizes assessing patient condition through non-invasive recordings, tailoring the model to the unique EEG patterns of each subject.

### Classification of MCI with EEG features

The model lied at the core of a clinical pipeline to determine the severity of each patient’s condition through a quantitative analysis of their resting EEG signal. The EEG signal was processed through the model to extract more informative biomarkers than standard EEG measures. We used as benchmark for EEG performances in classification of MCI state algorithms based on spectral and connectivity features, which are the most common tool to assess MCI conditions from EEG [11, 45-52] and are even investigated as possible markers of progression to AD [37]. The final result was in line with previous studies performing MCI classification with these features [9, 10, 13, 14, 53] but was vastly outperformed by the model-based procedure (Figure 6).

However, the range of possible EEG features extend beyond the ones we tested. Promising results have been achieved through network [54] or microstate analysis [13, 55]. Moreover, EEG features collected during cognitive tasks are likely to be more informative than those acquired at rest and hence lead to a more efficient classification EEG synchronization in MCI and AD (56). A potential alternative approach is automatic learning of features based on deep learning tools [57]. Novel approaches can improve not only classification with EEG features, but also improve the design personalized models. Previous works utilized model personalized with structural scans as [19] to derive novel features to support classification of AD and MCI patients [53,58]. Beside the difference in the approach highlighted in the previous section, we also note that in our case network model features replace features based on recordings in the classification algorithm, rather than being combined with them, nonetheless enabling us to obtain a much higher accuracy in discriminating healthy and MCI subject.

High performance MCI classification based solely on EEG analysis could greatly enhance the manageability of the disease in disadvantaged areas enabling clinicians to obtain a reliable diagnosis with affordable means. Harnessing the precision of EEG analysis for classification could support disease management in resource-limited regions grappling with a dearth of MRI or PET machines that nowadays impedes reliable diagnoses.

### Model prediction of structural alterations and clinical outcome

Detecting cognitive decline at its nascent stages is pivotal, as it offers a window of opportunity for early therapeutic interventions. However, the complexity of diagnosing SCD arises from its elusive and heterogeneous nature, characterized by the absence of distinctive symptomatology [3]. Unlike clinical dementia, SCD lacks the objective cognitive deficits that can be readily quantified through standardized neuropsychological assessments, rendering diagnosis reliant on subjective reports of cognitive decline from individuals themselves or their informants.

This inherent subjectivity in symptom reporting poses challenges in ascertaining the validity and specificity of SCD as a clinical entity. Our model-based approach was able to partition SCD into two subsets with different CSF biomarkers (currently the most reliable biomarker of AD [20] (Figure 5G). The prediction of CSF biomarkers in SCD subjects was never attempted before to the best of our knowledge, and in fact was unachievable when using standard quantitative EEG analysis.

(Figure 2F). This constitutes the first example of prediction of biological hallmarks of AD from EEG recordings, a fortiori in preclinical subjects without evident symptomatology. Moreover, all the three SCD subjects that converted in the year following the recordings were classified at risk of conversion by our model. These results pave the way for a novel approach to EEG analysis, that could extract from such an affordable and non-invasive technique information until now obtainable only via more complex and/or stressful exams. It would be extremely beneficial, for both medical facilities and patients, to be able to predict the state of structural alterations without having to perform invasive procedures such as the lumbar puncture to assess the presence of biological hallmarks of such alterations.

### Limitations and future directions

A first limitation of our study pertains to the size of the three participant groups: in particular the smaller size for the control group compared to the other two groups. We ensured to minimize the bias introduced by this discrepancy in the statistical analysis (see Methods). Furthermore, the cerebrospinal fluid (CSF) was analysed only on a limited subset of subjects.

Another limitation lies in the selection of the regions of interest that exhibit impairment. While our work focuses on predicting structural changes linked to AD in the absence of direct structural data, we had to draw from existing literature [20] to determine the regions of interest, referring to the well-known Braak stages [39] (see Methods). This implies that our findings might be influenced by the choice of the impaired regions. To address this, we explored various combinations of impaired regions (based on Abeta and Tau Braak stages, their combination, intersection, or solely the Hippocampus), revealing ABeta Braak stages as the optimal choice; these changes did not qualitatively affect the results (data not shown).

Third, the aggregation of meso-scale and macro-scale parameters, while facilitating precise model customization and severity assessment, necessitates nuanced assumptions. Specifically, the proportionality between neuroplastic adaptations (represented in the model as increased short-range structural connectivity in impaired regions) and white matter degradation (manifested as a loss of long-range structural connectivity) could be highly subject-dependent [35, 36]. While we experimented with diverse ratios of these quantities to identify the most suitable ratio for our dataset (finally settling for the 1:1 ratio), the inherent subjectivity of deviation from this aggregated value remains unaddressed in this study.

These highlighted challenges delineate promising avenues for future research. One possible direction involves enhancing model realism by integrating structural data from PET or MRI scans into the pipeline, enabling improved identification of impaired regions and quantification of white matter atrophy and neuroplasticity ratios, which significantly influence the model’s structural alterations. Longitudinal follow-up studies on the enrolled subjects, already planned, will assess the strength of the model’s predictive capacity. Finally, the model could be validated using EEG recordings during tasks that assess cognitive impairments by adding an extension able to capture the effect of visual stimuli.

Advancing the model’s power and versatility could involve co-simulations with software like NEST [59] or ANNarchy [60], mirroring prior applications in the study of deep brain stimulation effects in Parkinson’s disease [61]. These multi-scale simulations can encompass alterations spanning from microscopic phenomena such as neuron loss in regions like the hippocampus [62] to the meso- and macroscopic effects discussed in our study [15].

In conclusion, our investigation proposes an innovative path where computational and quantitative EEG analyses converge, extracting pivotal clinical markers and surpassing traditional approaches. The proposed pipeline not only enables the quantification of disease severity through structural insights inferred with non-invasive methods, but also facilitates the prognosis of disease progression, even in subjects in preclinical stages of the disease who lack evident symptomatology. This novel approach lays the groundwork for a more affordable and effective diagnosis of neurodegenerative disorders. It holds potential for further evolution through the inclusion of structural imaging modalities and multiscale co-simulations, paving the way for clinical investigations integrating non-invasive biomarkers with computational frameworks, promising rapid and precise monitoring of neurodegenerative conditions.

## Data Availability

All data produced in the present study are available upon reasonable request to the authors

## Supplementary information

Supplementary tables and figures can be found in the attached Supplementary Materials file.

## Acknowledgements

We thank dr. Sara Moccia and Angelo Lasala for precious hints on the classification pipeline.

## Data and Code Availability

Raw EEG data are available upon request for research and scientific purposes. All the code used to perform the simulation and analyses hereby discussed is implemented in Python 3.27. The EEG preprocessing was conducted with the *EEGLAB* Matlab Toolbox. We used the Python packages *Scipy* to perform the statistical analyses, *MNE* to compute EEG features, *Sklearn* for the machine learning pipeline, and The Virtual Brain software (*tvb* Python library) to build the model and simulate brain activity. Code is available upon request.

## Consent Statement

We state that every subject enrolled in this study provided informed consent prior to participation.

