## Supplementary Materials for "Personalized modeling of neurodegeneration determines dementia severity from EEG recordings"

**Supplementary Material**

### Supplementary Methods

### Subjects recruitment

As part of a longitudinal, clinical–neuropsychological–genetic survey on SCD and MCI, we included 102 consecutive spontaneous patients who self-referred to the Centre for Alzheimer’s disease and Adult Cognitive Disorders of the Careggi Hospital in Florence. All the patients were Caucasian. Inclusion and exclusion criteria were described in a previous work by our group. All participants underwent an extensive neuropsychological battery, assessment of cognitive complaints, and peripheral blood collection to analyze Apolipoprotein E (APOE), HTT, and BDNF genotypes. We divided our sample into two groups: n = 58 patients classified as SCD, according to the terminology proposed by the Subjective Cognitive Decline Initiative (SCD-I) Working Group (i.e., presence of a self-experienced persistent decline in cognitive capacities with normal performance on standardized cognitive tests). The other patients (n = 44) were instead classified as MCI, according to the (NIA-AA) criteria for the diagnosis of MCI. In addition, we included a group of n = 17 healthy, age-paired subjects as a control group. The local ethics committee ap- proved the protocol of the study. All participants gave written informed consent. All procedures involving experiments on human subjects were done in accordance with the ethical standards of the Committee on Human Experimentation of the institution in which the experiments were done or in accordance with the Helsinki Declaration of 1975. Specific national laws have been observed.

#### EEG preprocessing and feature extraction

In order to discard artifacts (electrophysiological and non) from the raw EEG signals, we run a custom pre-processing pipeline written in Matlab with the use of the EEGLAB toolbox functions (*1*). The preprocessing pipeline consisted in the PREP pipeline (*2*) followed by ICA removal of artifactual components (*3*). The first step of preprocessing included the use of PREP pipeline, which performs several preprocessing steps automatically, allowing to obtain robust average re-referenced signals. Initially, PREP high-pass filters signals of all channels, by means of a Hamming windowed FIR filter (using EEGLAB’s pop_eegfiltnew function) with a 1 Hz cut-off frequency. Line noise at 50 Hz and its harmonics were removed by using the CleanLine EEGLAB plugin. Noisy channels, i.e. those channels having abnormal and/or uncorrelated activity compared to others were removed by using PREP noisy channel subroutine, which performs the bad channel selection by combining an ensemble of methods: the deviation criterion, the correlation criterion, the noisiness criterion and the predictability criterion. Remaining channels activity was used to estimate a robust average reference, based on robust statistics such as the median and interquartile range. Finally, removed channels were interpolated, by means of spherical interpolation. The obtained re-referenced and filtered signals were then subjected to the second preprocessing step. Independent components were extracted by using the Infomax ICA algorithm, as implemented in binica EEGLAB routine. A semi-automated procedure was then used to distinguish between brain-related components and artifactual ones. We used ICLabel to classify automatically independent components into brain or artifactual components (line noise, muscle, eye, channel noise, heart, “other”) based on a neural network trained on crowd-sourced data. ICLabel returns the probability of each component to belong to one of the above-mentioned classes. We then used DIPFIT to perform a single dipole fitting of the independent component map onto a template brain (MNI-152 atlas). Given that brain components should be dipolar, a high residual variance of the fitted dipole should indicate a low probability of the component being brain related. Hence, components labeled by ICLabel as “brain” with a confidence higher than a set threshold (we used 75%) and having fitted dipole residual variance lower than another threshold (we used 20%) were retained in the final signals. Noise components with high confidence and high dipole residual variance were instead automatically removed from the ICs list. All the remaining components were inspected visually and flagged either as brain or non-brain depending on their power spectra profiles and time-courses. Channel-level signals were finally reconstructed from the reduced IC space, only including brain-related sources. Finally, we performed a visual inspection of the cleaned signals, to remove possible remaining artifacts (e.g., temporally localized muscle activity not removed by the ICA procedure). Prior to any quantitative analysis, signals were bandpassed in the range 0.5-45 Hz by using a Butterworth filter of first order (*4*). Feature values were divided into N equipopulated bins, where N is given by the dimensionality of the dataset (N = 119) and the bin width is determined by the cumulative distance of each point from its *k*-nearest neighbours (*5*) with *k* = 10. Bias correction was performed by means of quadratic extrapolation (*6*). The significance of the given mutual information was computed by iterating 100 times a bootstrap test, thus generating a null distribution. We defined the significance threshold on the basis of the so-computed mutual information by taking the 95th higher value obtained by computing Eq. 1 in the bootstrapped distributions (p < 0.05).

### Supplementary Figures and Tables

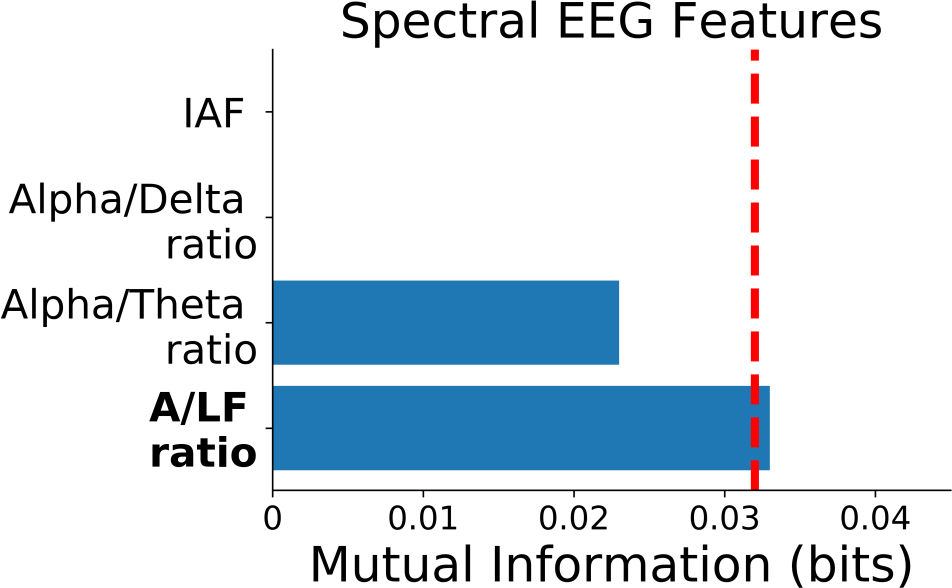
**a**

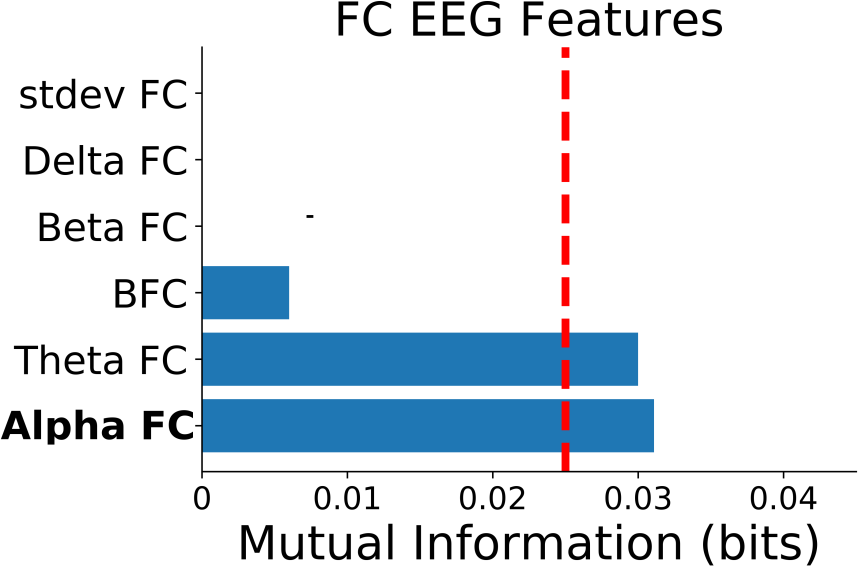
**b**

Supplementary Figure 1: EEG features selection **a**: Information carried by spectral features about subjects’ condition **b**: Information carried by FC features about subjects’ condition. Vertical red lines represent the significance threshold (p < 0.05) for the most informative feature, determined by bootstrap technique (see main text)

| **Parameters** | **Classifier** | **Accuracy** | **F1 score** |
| --- | --- | --- | --- |
| A/LF ratio - FC alpha band average | SVM, rbf kernel | 0.68 | 0.73 |
| A/LF ratio - FC Alpha band average | Decision Tree | 0.66 | 0.67 |
| A/LF ratio - FC Alpha band average | QDA | 0.60 | 0.69 |
| A/LF ratio - FC Alpha band average | Naive Bayes | 0.57 | 0.67 |

Supplementary Table 1: Top classifier performances trained with combinations of the most informative EEG spectral and FC features. Note that all performances are inferior compared to those of the Random Forest algorithm that we eventually preferred.

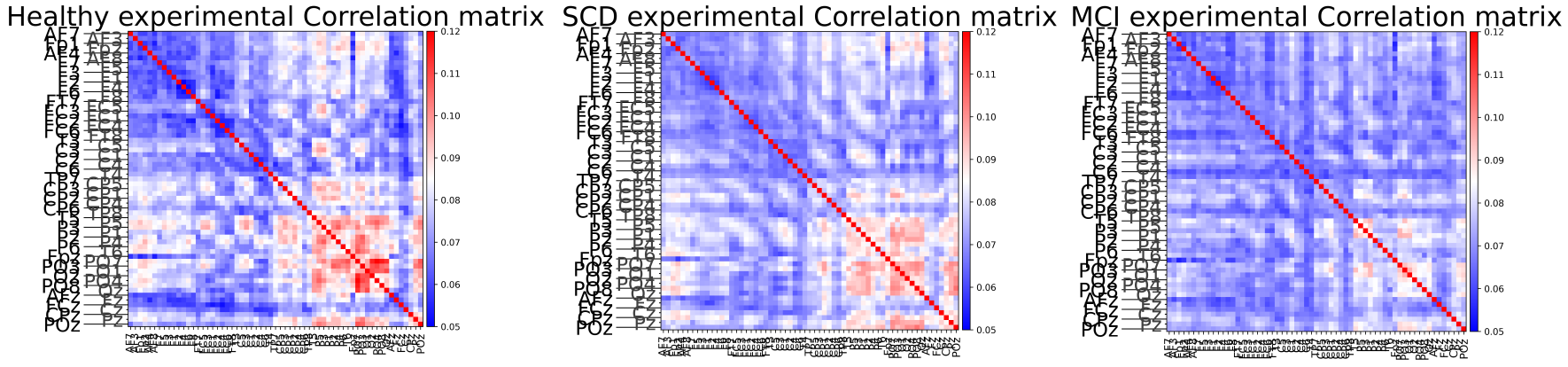

## a

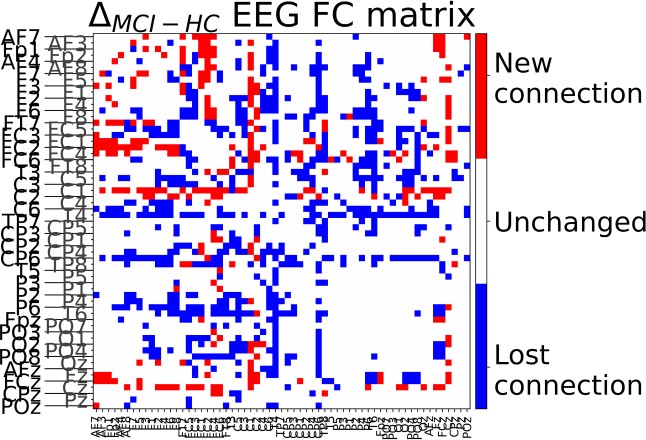

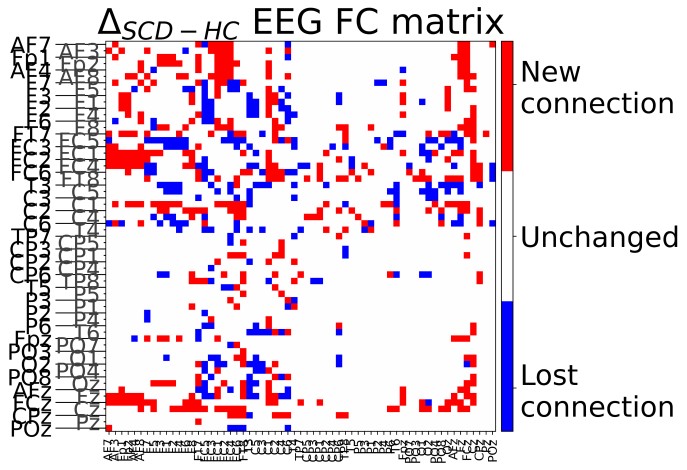

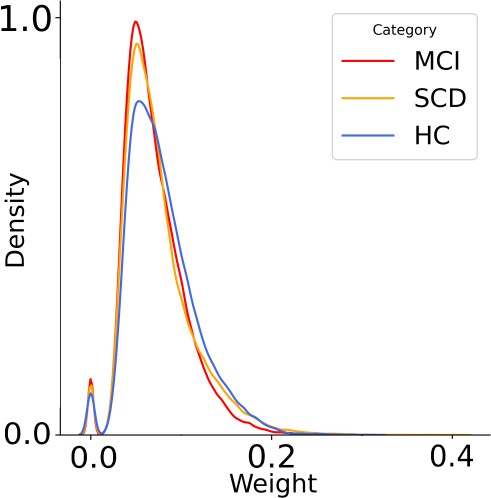

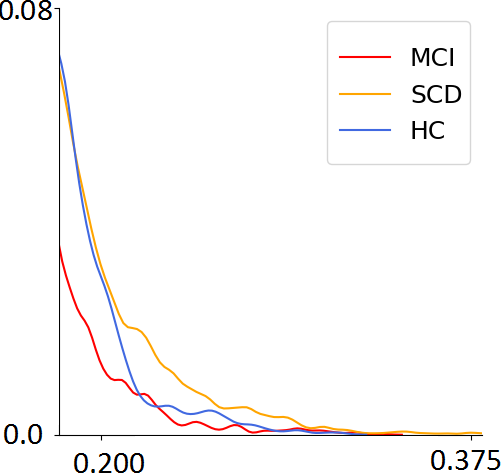

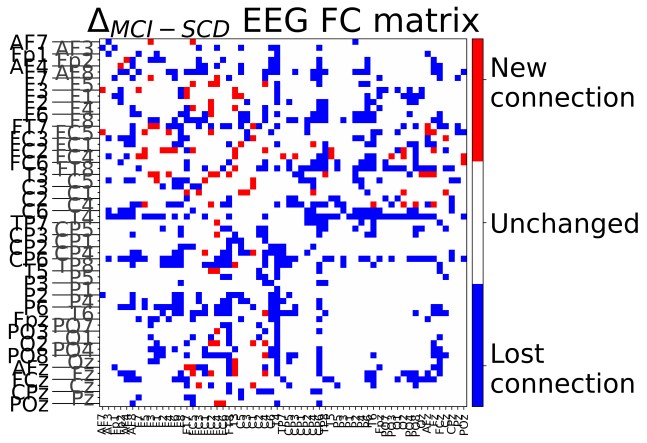

**b**

**c**

Supplementary Figure 2: **a**: Functional connectivity matrix averaged over subjects of the different groups **b**: Distribution of functional connection weights per each group. Inset shows that high connective values are more common in SCD subjects than in HC and MCI **c**: Differences between connectivity matrices displayed in (a).

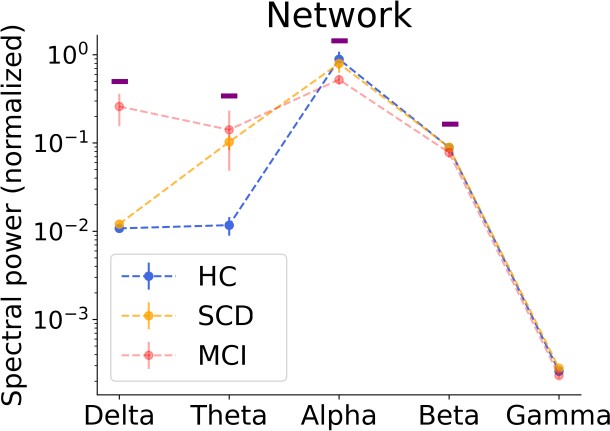
**a	b**

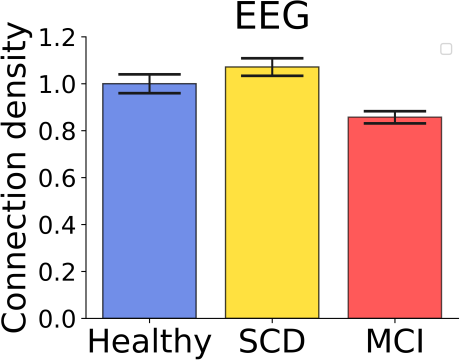

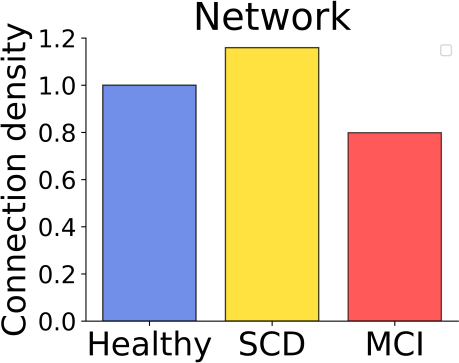

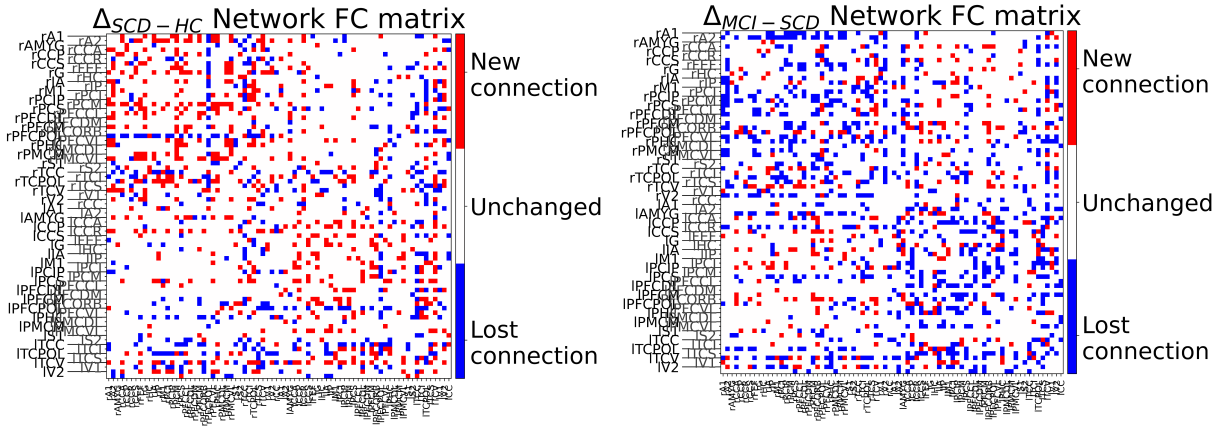

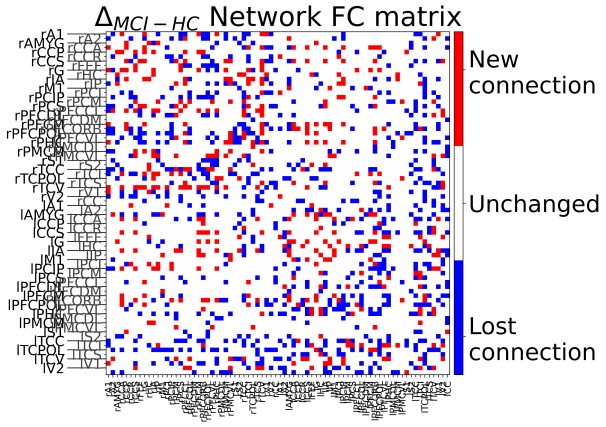

**c**

Supplementary Figure 3: EEG connectivity analysis **a**: FC Delta matrices computed from simulated signals. Color code is the same as in 2. **b**: Barplot of the number of significant connections for each population in both empirical and simulated case, normalized to the healthy value. Y axes not in scale. Note that error bars are not reported for quantities computed from simulated signals, as these are highly stable across multiple runs of the code (error less than 10*−*5 of the mean value) **c**: Normalized power spectra bands for simulated signals. Purple bands stand for frequency bands in which the model shows the same behaviour observed in the experimental case (same ordering between populations).

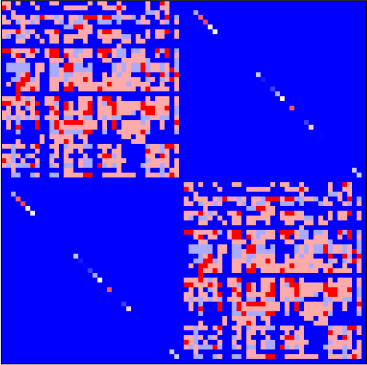

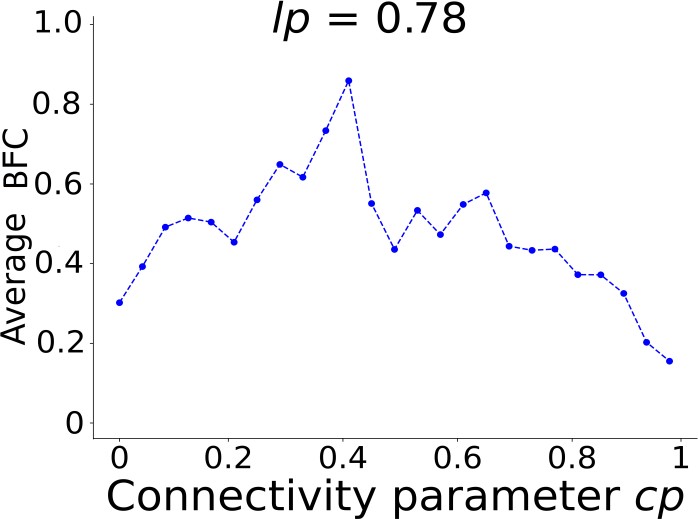

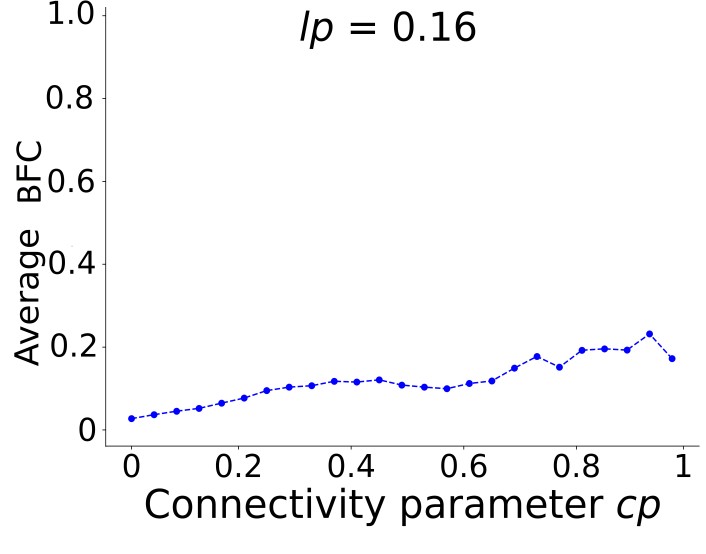

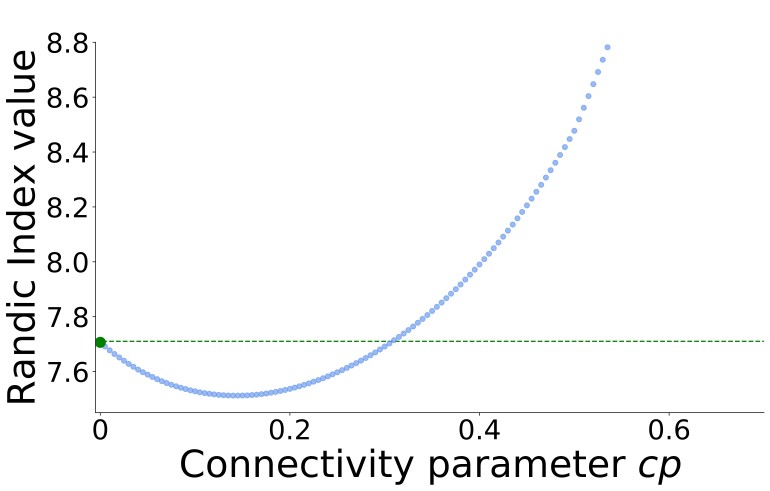

**a**

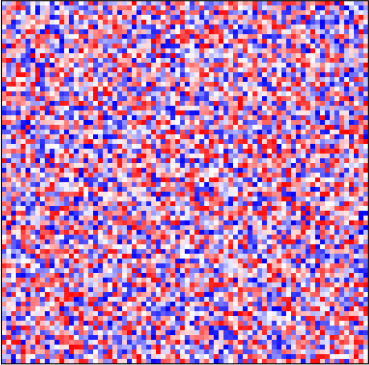

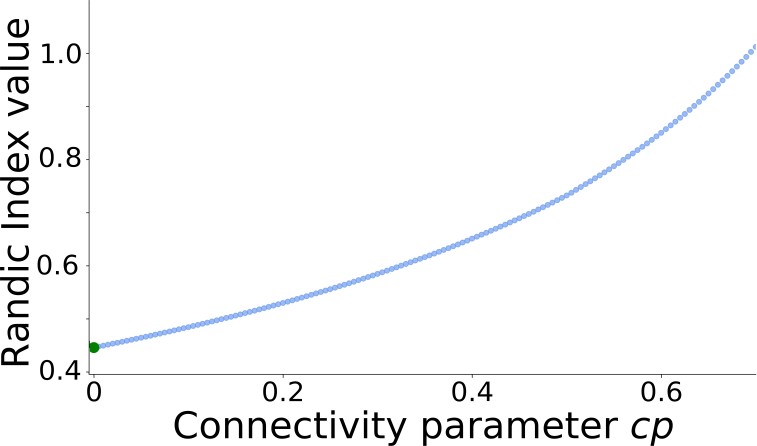

**b**

Supplementary Figure 4: **a**: FC and Randic index as a function of *cp* for the model implemented with the standard TVB structural connectivity matrix. **b**: FC and Randic index as a function of *cp* for the model implemented with a random structural connectivity matrix. Note how the nonlinearities are absent in the random case, suggesting how the non-trivial topology of the network is a key-ingredient for the non- linearities observed both experimentally and in the model simulations.

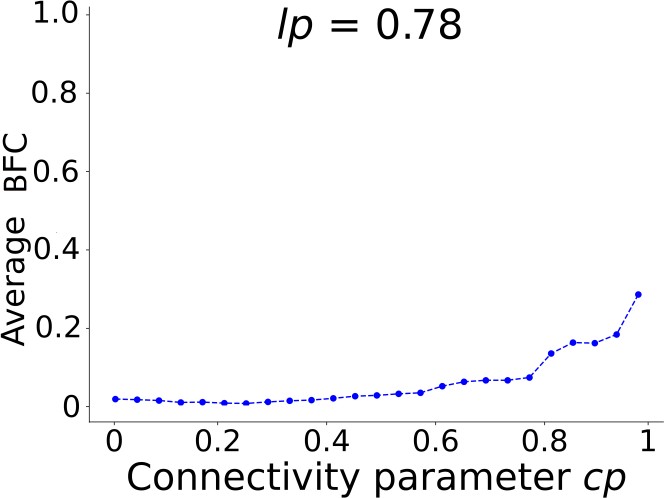

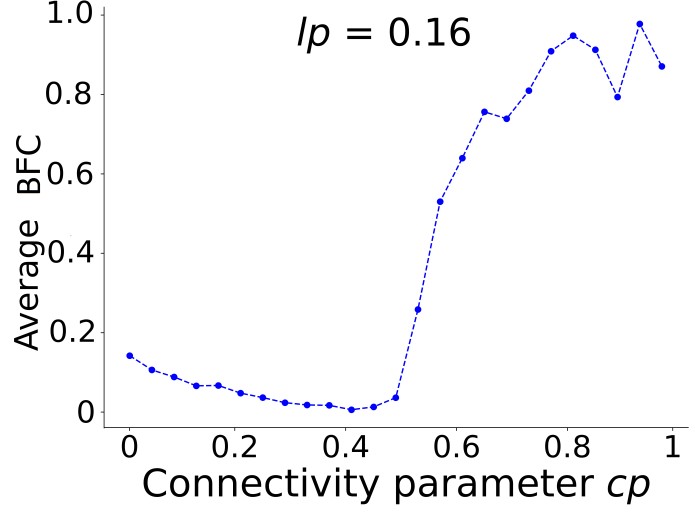

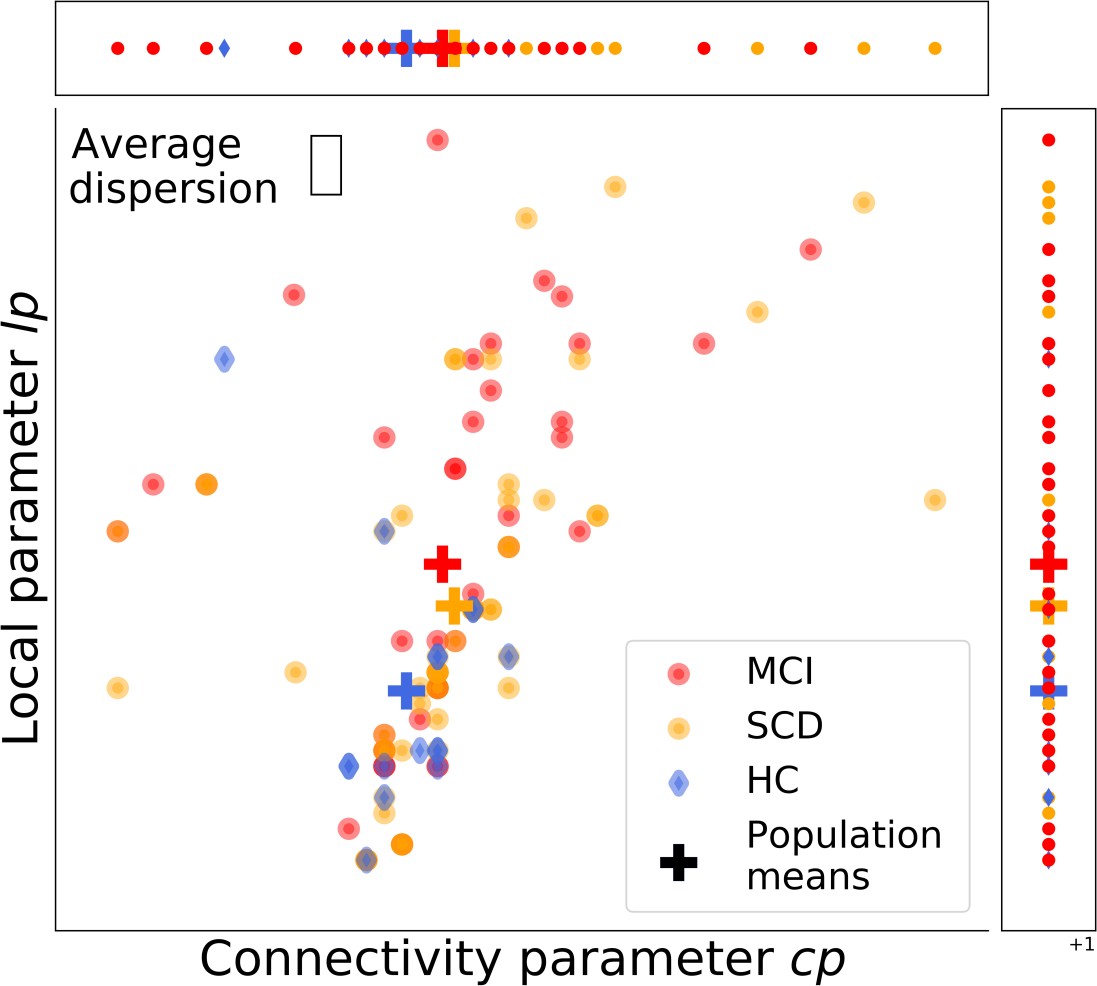

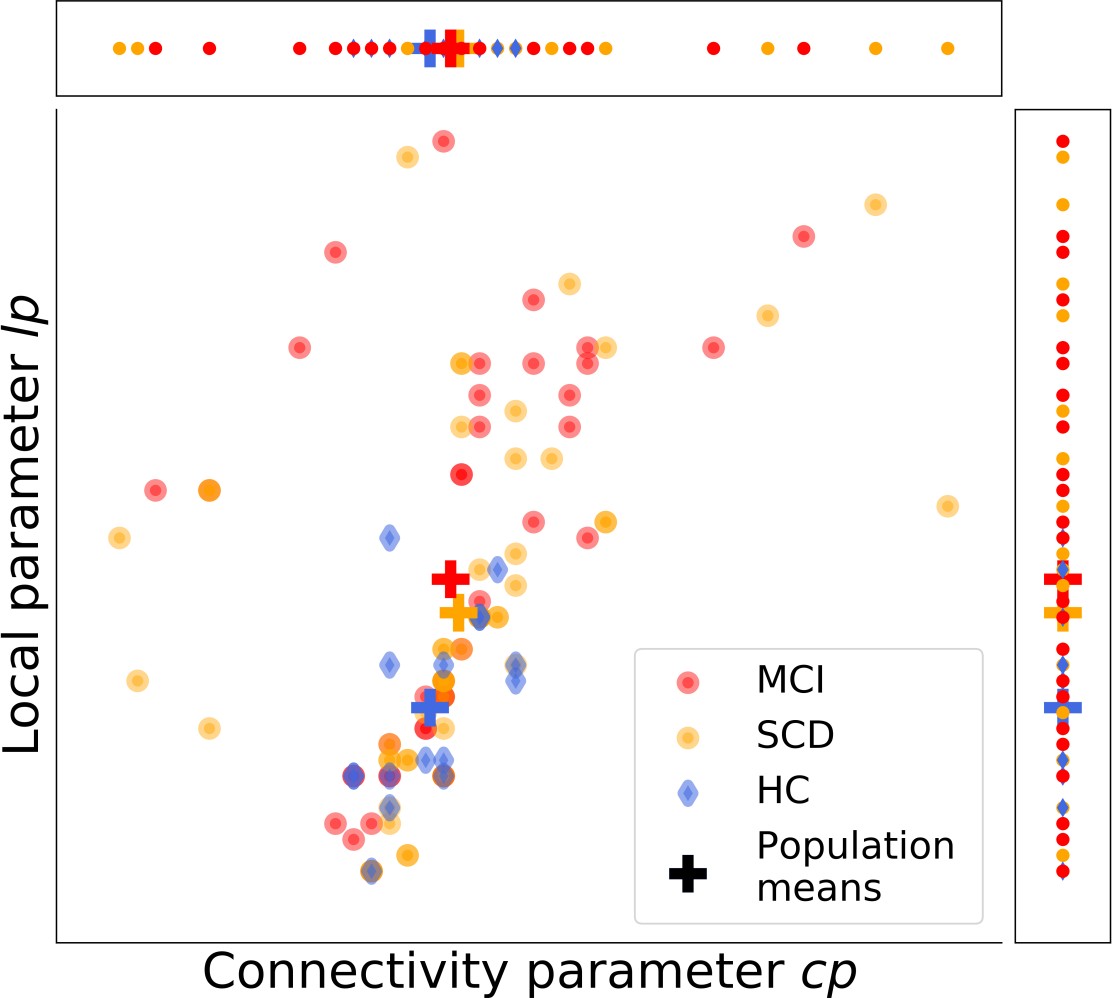
 **a	b**

Supplementary Figure 5: Robustness test for the subject-dependent parameters determination **a**: Distribution of best fitting parameters for each subject as shown in Figure 5. Parameters were determined subject-wise from the values of BFC and A/LF ratio. **b**: Distribution of best fitting parameters for each subjects, determined from the values of BFC and A/LF ratio after a gaussian random shift of the experimental values.
